# Integrating Causal Inference into Pharmacovigilance: Target Trial Emulations for Proactive Signal Detection of Atorvastatin Initiation in Medicare Beneficiaries

**DOI:** 10.64898/2026.07.01.26356874

**Authors:** Christopher G. Rowan, John Tazare, Macarius M. Donneyong, Minh Tran, Shivani Srivastava, Nancy A. Dreyer, Camille Maringe

**Author notes:** **Corresponding author:** Christopher G. Rowan, PhD, 4030 Calle Marlena, San Clemente, CA 92672, https://www.veritasrf.org/.

## Abstract

**Importance:** Adverse drug events (ADEs) in older adults are a substantial public health burden, yet spontaneous reporting systems detect them poorly owing to underreporting and the lack of a defined population. These limitations are of particular concern for older adults, who are underrepresented in pre-approval trials yet at elevated ADE risk owing to polypharmacy, multimorbidity, and age-related changes in drug metabolism.

**Objective:** To develop and apply an active, claims-based pharmacovigilance framework using sequential target trial emulation to detect ADE signals in older adults, with atorvastatin as the initial application.

**Methods:** Using Medicare fee-for-service claims (2017-2019), we studied statin-naïve beneficiaries aged 65 years or older following hospitalization for myocardial or cerebral infarction. We emulated up to 14 daily sequential trials from the discharge date, classifying patients as initiating atorvastatin (A1), initiating a different medication (A2), or no new medication (A0); the primary contrast was A1 versus A2. For each trial, incident ADEs were ascertained and classified into 552 outcomes based on the Clinical Classifications Software Refined categories. Per-protocol effect estimates were measured over a 6-month follow-up period using Fine-Gray regression weighted by the inverse probability of treatment (IPT) and censoring, treating death as a competing risk, with the false discovery rate controlled via the Benjamini-Hochberg procedure. A signal was declared when the q-value was ≤ 0.10 and the subdistribution hazard ratio (sHR) was ≥ 1.20 in any prespecified analytic stratum (sensitivity analyses used thresholds of q ≤ 0.20 and sHR ≥ 1.20).

**Results:** Of 70,130 eligible patients, 39,948 initiated atorvastatin (A1), 19,182 initiated another new medication (A2); after IPT-weighting, baseline characteristics were closely balanced. After excluding outcomes with sparse cell counts, 295 outcomes were analyzed; five met the primary signal detection criteria: valve disorders (sHR 1.71, 1.20-2.43); sprains and strains (sHR 1.79, 1.26-2.54); general sensation/perception symptoms (sHR 1.23, 95% CI 1.11-1.36); abnormal findings without diagnosis (sHR 1.55, 1.18-2.05); and prediabetes (sHR 1.71, 1.24-2.36). In the sensitivity analysis, we additionally detected: posthemorrhagic anemia, hemorrhagic stroke, varicose veins, other circulatory and skin conditions.

**Conclusions:** A claims-based framework using sequential target trial emulations detected both expected and previously unrecognized ADE signals following atorvastatin initiation in older adults, offering a systematic alternative to passive surveillance that can be extended to other commonly prescribed medications. Confirmatory analyses with disaggregated outcomes and narrower comparators are underway and will be reported separately.

**KEY POINTS:** *Question:* Can an active, claims-based pharmacovigilance framework using sequential target trial emulation detect adverse drug event signals among older Medicare beneficiaries who initiated atorvastatin?

*Findings:* In this cohort study of 59,130 Medicare beneficiaries discharged after myocardial or cerebral infarction, a hypothesis-free scan across hundreds of prespecified outcomes detected five primary signals (valve disorders, sprains and strains, sensory symptoms including dizziness, abnormal laboratory findings, and prediabetes) and additional signals in sensitivity analyses (including hemorrhagic stroke), several of which align with known or labeled statin adverse effects (serving as positive controls that validate the framework) while others remain uncertain.

*Meaning:* This framework offers a systematic, quantitative, and proactive alternative to spontaneous reporting for detecting medication safety signals in high-risk older adults.

**PLAIN LANGUAGE SUMMARY:** Older adults use more prescription drugs than any other age group, yet they are often left out of clinical trials conducted before a drug is approved. As a result, the safety of many medications in older patients is poorly understood until the drugs are already in wide use. The main system for detecting new drug safety problems in the United States, the FDA Adverse Event Reporting System, relies on voluntary reports and captures only a small fraction of harms, which limits its usefulness.

Using Medicare reimbursement records, the researchers developed an active monitoring method and tested it on a commonly used cholesterol-lowering medication (atorvastatin) – that is frequently prescribed after a heart attack or stroke. They followed thousands of patients for six months, statistically balancing those who started atorvastatin with those who started a different new drug, and screened for hundreds of possible side effects. Evidence of potential medication harm emerged along a clear range of how likely the drug caused each problem. This ranged from well-known side effects (such as diabetes and sprains and strains) and outcomes already noted in the product labeling or prior research (such as hemorrhagic stroke and dizziness) to more uncertain associations (such as cardiac valve disorders) that are difficult to interpret. Overall, atorvastatin appeared largely safe in this older population, and the same method can now be used to study other widely used drugs.

## INTRODUCTION

Pharmacovigilance is the scientific discipline concerned with the detection, quantification, and contextualization of adverse drug events (ADEs) after regulatory approval.^1^ Preapproval randomized trials often lack statistical power to detect uncommon events, have limited follow-up, and enroll participants who may appreciably differ from those who will use the medication in routine clinical practice.^2–9^ These limitations are particularly consequential for older patients, who bear a high burden of multimorbidity and polypharmacy and who experience age-related changes in pharmacokinetics and pharmacodynamics that may alter drug metabolism and increase susceptibility to ADEs. Older adults are also frequently underrepresented in preapproval trials; thus, reliable evidence on real-world ADEs in this population remains limited.^7,10–19^

Conventional pharmacovigilance relies primarily on passive surveillance systems, such as the FDA Adverse Event Reporting System (FAERS). These systems depend on voluntary or “spontaneous” reports from clinicians, patients, and manufacturers. Although they have identified important safety signals, these anecdotal reports suffer from substantial underreporting, variable report quality, and the absence of a defined population denominator.^20^ In the absence of a known denominator of treated patients, incidence rates cannot be estimated, and quantitative risk assessment remains imprecise. These shortcomings limit the ability of traditional methods to reliably detect, characterize, and contextualize suspected ADEs, especially among older patients with both heightened ADE risk and therapeutic need.^9–11,21–31^ While the FDA’s Sentinel initiative represents an advancement in assessing potential medication-related harm, it primarily conducts reactive, FDA-directed investigations and is not specifically focused on older adults or proactive pharmacovigilance.

To address these limitations and build upon established safety evidence pertaining to atorvastatin in routine clinical practice, we used population-based longitudinal Medicare claims data to proactively emulate a series of pragmatic target trials as a high-dimensional, hypothesis-free ADE signal detection study, with hospital discharge after myocardial or cerebral infarction serving as the empirically defined time zero event.^32–34^ Atorvastatin was selected as the initial application both because of its widespread post-discharge use in this population and because its well-documented safety profile provides positive controls to rigorously evaluate the framework’s ability to recover known signals while proactively screening for unknown or poorly characterized ADEs. Among older, statin-naïve Medicare beneficiaries who met all eligibility criteria, we asked the following causal question: *What is the per-protocol causal effect of initiating atorvastatin versus initiating another new outpatient medication after hospitalization for myocardial or cerebral infarction —both in addition to other newly initiated or continued medications—on the subdistribution hazard ratio of each of hundreds of potential ADEs, treating death as a competing risk?*

## METHODS

### Study Population and Sequential Trial Framework

The study population included Medicare fee-for-service beneficiaries aged 65 years or older who were statin-naïve prior to hospitalization for myocardial infarction or cerebral infarction (length of stay ≥ 3 days) and discharged home during the study period (January 1, 2017 to January 31, 2019). We refer to this hospitalization and subsequent discharge as the time zero event, an index event identified empirically in prior research among older statin-naïve Medicare beneficiaries.^34^

The discharge date marked the origin of a 14-day sequence of daily trials (trials 0 through 13), with the discharge date serving as the trial 0 start date. Upon initiation of each trial, eligibility was assessed using data from the preceding 12 months (baseline period). Eligibility for each trial required that the beneficiary be alive, statin-free, continuously enrolled in Medicare Parts A, B, and D, free of the outcome under study, and without a contraindication to atorvastatin (acute liver disease, cirrhosis, or hypersensitivity to atorvastatin).

These criteria identified beneficiaries eligible for post-discharge pharmacotherapy in the absence of recent statin exposure or clear contraindications. Application of these criteria at each trial start date aligned the assessment of eligibility, the assignment of treatment strategies, and the start of follow-up with the protocol of the hypothetical target trial that we sought to emulate (supplemental material **Table S6**).

### Treatment Strategies

Treatment strategies were defined and classified according to observed pharmacy dispensing claims on each trial start date. We emulated three pragmatic treatment strategies to reflect real-world clinical decision-making at the point of potential treatment initiation following an acute event: (A1) initiation of atorvastatin in addition to other prevalent and incident medications; (A2) initiation of a new outpatient medication other than atorvastatin in addition to other prevalent and incident medications; and (A0) no initiation of a new medication.

Eligible beneficiaries whose observed medication dispensing data were consistent with strategy A1 or A2 in a given trial were included in that trial (as A1 or A2) and rendered ineligible for all subsequent trials. Beneficiaries classified under strategy A0 in a given trial remained eligible for subsequent trials until they initiated either strategy A1 or A2. This sequential daily trial design aligned each trial start date with the observable clinical decision point for treatment initiation. By defining eligibility, assigning treatment strategies, and initiating follow-up concurrently within each trial—and by excluding from later trials those who had initiated treatment—we eliminated immortal time bias that would otherwise arise in a single-trial design in which eligibility was classified on the date of medication initiation.

### Follow-up and Censoring

In each trial, follow-up began the day after the trial start date and continued for 182 days or until the outcome under study or a censoring event occurred. Each trial start date was used to ascertain baseline characteristics, exclude beneficiaries with prior evidence of the outcome under study, confirm eligibility, and classify treatment strategy assignment. Beneficiaries who experienced the outcome of interest on the trial start date were excluded from that trial and all subsequent trials, although they could have been included in earlier trials. The one-day delay in the start of follow-up ensured that outcome ascertainment began after determination of eligibility and treatment assignment.

Censoring was classified as informative (death), administrative (end of the study period or loss of Medicare Parts A, B, or D coverage), or artificial (deviation from the assigned treatment strategy, used to estimate the per-protocol effect). Deviation from the assigned treatment strategy was defined as the earliest of initiation of a statin other than atorvastatin, switching between treatment strategies (i.e., A1 to A2 or A2 to A1), or discontinuation (treatment dispensing gap exceeding 45 days). For beneficiaries assigned to strategy A2, one of the medications newly initiated on the trial start date was selected at random to classify discontinuation (see supplemental material **Table S1** for the frequency distribution of randomly selected A2 medications). Upon discontinuation of strategy A1 or A2, 30 days were added to the days supplied to accommodate imperfect adherence. To ensure symmetric handling of artificial censoring due to switching, switches from strategy A1 to A2 were randomly assigned according to the observed distribution of switching from A2 to A1, reflecting the heterogeneous composition of medications in strategy A2.

### Outcomes

We generated a broad array of outcomes as potential ADE signals from International Classification of Diseases, Tenth Revision, Clinical Modification (ICD-10-CM) diagnosis codes recorded during routine care in Medicare claims. We mapped codes from inpatient, emergency department, and outpatient claims during follow-up to the Healthcare Cost and Utilization Project Clinical Classifications Software Refined (CCSR) categories (outcomes). The CCSR system aggregates all ICD-10-CM codes into 552 clinically coherent groups across 22 organ systems. For each category, an incident outcome was defined by the absence of that outcome in the year before the trial start date (inclusive of the trial start date) together with at least one qualifying code during follow-up; only the first outcome occurrence was analyzed. Baseline evidence of the outcome required one code from an inpatient or emergency department claim or two codes on separate dates from outpatient claims. For each of the outcomes considered and each of up to 14 sequential trials (for each outcome), cohorts were redefined by excluding patients with prevalent outcomes in the baseline year and by reassessing eligibility at every daily trial origin; composition of each trial for each outcome therefore varied by timing of treatment initiation and outcome-specific criteria.

### Baseline Covariates

Baseline covariates were ascertained during the year preceding each trial start date (inclusive of the trial start date unless otherwise specified). Demographic characteristics (age, sex, race, dual eligibility, and disability status) were obtained from the Beneficiary Summary File. Healthcare utilization was quantified by the number of inpatient admissions, inpatient days, emergency department visits, and the total number of unique service dates; total medical costs were calculated over the same interval.

Comorbidities were ascertained from inpatient, emergency department, and outpatient claims using ICD-10-CM codes mapped to CCSR categories. One code sufficed for inpatient or emergency department diagnoses; outpatient diagnoses required two codes on separate dates. The Charlson Comorbidity Index was computed using a weighted algorithm. Baseline pharmacotherapy was identified from Part D dispensing claims (excluding the trial start date) using generic names and GCDF codes linked to the First Databank and American Hospital Formulary System classifications, yielding 417 baseline medication variables across 20 major classes and 397 subclasses. Baseline inpatient procedures were classified from hospital claims using ICD-10-PCS codes into 320 CCSR categories across 31 domains. Outpatient procedures from facility claims were classified using Current Procedural Terminology (CPT) and Healthcare Common Procedure Coding System (HCPCS) codes and mapped to 245 CCSR procedure categories.

### Statistical Analysis

For each outcome under study, we emulated sequential target trials among statin-naïve beneficiaries. The primary exposure contrast of interest was strategy A1 (initiation of atorvastatin) versus strategy A2 (initiation of any other new outpatient medication). Treatment strategy A0 was retained to preserve the structural integrity of the sequential trial design but was excluded from the primary exposure contrast. Beneficiaries who initiated no new medication following a hospitalization for myocardial infarction or cerebral infarction (A0) likely differed in important prognostic ways from those who initiated new pharmacotherapy; we therefore did not analyze A0 to mitigate channeling bias.

Outcomes with fewer than 70 total events, outcomes representing previously diagnosed conditions [e.g., Sequela of hemorrhagic cerebrovascular disease (CIR022)], and the prespecified lipid metabolism outcome (CCSR END010) directly related to the atorvastatin indication were excluded from the analysis. For each remaining outcome, sequential trials were retained until either arm (A1 or A2) contained fewer than 40 beneficiaries; individuals with baseline evidence of the outcome under study were excluded from that trial and all subsequent trials, and same-day outcome and death events were classified as outcomes.

Within each trial for a given outcome, stabilized inverse probability of treatment weights (IPTW) were estimated using logistic regression. Stabilized inverse probability of censoring weights (IPCW) for treatment strategy deviation were estimated using Cox proportional hazards regression to account for the time-to-event nature of artificial censoring. Covariate selection for both models was performed adaptively and independently per trial by iterative removal of variables with zero variance, perfect prediction, Pearson correlation exceeding 0.95, or cell counts below 20. The product of the stabilized weights (IPTW*IPCW), truncated at 10, was applied to data stacked across retained trials for each outcome.^35–37^

The per-protocol effect of atorvastatin initiation (A1) versus initiation of any other new medication (A2) was estimated as the subdistribution hazard ratio (sHR) and 95% confidence interval (CI) over 182 days or until censoring, using weighted (IPTW*IPCW) Fine-Gray regression that treated death as a competing risk and incorporated robust variance estimation clustered on patient identifier.^38^ From the first trial (trial 0) for each outcome, we reported the cumulative incidence under each treatment strategy (A1 and A2), the risk difference, and the number needed to harm (NNH). Diagnostics evaluated weight distributions and post-weighting covariate balance.

Prespecified stratified analyses were performed by follow-up interval (days 1–60, 61–120, 121– 182) to examine potential violations of the proportional hazards assumption and effect modification,^39–41^ and within subgroups defined by age (65–74, 75–84, ≥85 years), sex (male, female), and race/ethnicity (White, Black, Asian, Hispanic).

### ADE Signal Detection Methodology

We employed a high-dimensional, hypothesis-free analytic framework to detect ADE signals by systematically screening hundreds of predefined outcomes. Within each *a priori* specified analytic stratum—defined by the overall cohort, time-stratified intervals, and demographic subgroups—we applied the Benjamini-Hochberg procedure across all qualifying outcomes to control the false discovery rate at q ≤ 0.10.^42,43^ Within each stratum, unadjusted p-values from weighted Fine-Gray models were ranked in ascending order and transformed to adjusted q-values. This stratum-specific approach ensured that ADE signals arising in clinically important subgroups received the same consideration as those identified in the overall cohort. An ADE signal was declared when the adjusted q-value was ≤ 0.10 and the sHR was ≥ 1.20.^44^ This dual criterion prioritized ADE signals with substantive population-level impact and clinical relevance while reducing the likelihood of detecting spurious associations attributable to residual confounding or chance.

Within each stratum, we restricted the Benjamini-Hochberg procedure to outcomes with at least 20 events among patients initiating treatment strategy A1 and at least 20 events among patients initiating treatment strategy A2. This minimum-event threshold was chosen to avoid including analyses with extremely sparse cell counts in the multiplicity correction, a decision motivated by the need to preserve the statistical integrity of the false discovery rate control despite the exploratory nature of the study. We conducted a sensitivity analysis using a relaxed FDR threshold (q ≤ 0.20) while retaining the sHR threshold of ≥ 1.20.

All analyses were conducted in Stata version 18.0 (College Station, TX) with fixed random-number seeds to ensure reproducibility. This study was reported in accordance with the TARGET (TrAnsparent ReportinG of observational studies Emulating a Target trial; supplemental material **Table S5**).

## RESULTS

### Study Cohort

We identified 70,130 statin-naïve Medicare fee-for-service beneficiaries aged 65 years or older discharged home after hospitalization for myocardial or cerebral infarction who met eligibility criteria on the time zero event date (supplemental material **Figure S1**). Within the 14-day trial period, 11,000 (15.7%) initiated no new oral medication (strategy A0), 39,948 (57.0%) initiated atorvastatin (strategy A1), and 19,182 (27.4%) initiated another new medication (strategy A2). These counts reflect the overall cohort and the first initiation of A1 or A2 during the 14-day trial period; outcome-specific analyses excluded patients ineligible for each trial.

As expected in this per-protocol emulation of sequential target trials, follow-up time and reasons for censoring differed between treatment strategies before we accounted for artificial censoring using inverse probability of censoring weights (supplemental material **Table S2**). Among beneficiaries who initiated atorvastatin (A1) or another new medication (A2), median follow-up was 148 days (interquartile range, 60–182) and 32 days (interquartile range, 0–69), respectively. These differences arose mainly from higher rates of treatment strategy deviation among A2 initiators, including initiation of a statin other than atorvastatin (36% versus 5%) and discontinuation of the assigned strategy (27% versus 19%). We therefore applied inverse probability of censoring weights to obtain unbiased per-protocol effect estimates.

### Baseline Characteristics

**Table 1** shows inverse probability of treatment–weighted baseline characteristics at first treatment assignment for the 59,130 patients who initiated atorvastatin (A1) or another new medication (A2) within the 14-day trial period. The overall mean age was 77 years; 52% were women and 84% were White. The mean Charlson Comorbidity Index was 6.2, with high prevalence of hypertension (84%) and lipid disorders (69%). Acute myocardial infarction occurred in 66% and cerebral infarction in 37%. In the prior year, 21% had been hospitalized, 44% had an emergency department visit, and mean healthcare costs were approximately $26,400. Common medications and procedures were balanced between groups. These characteristics describe the overall population at first treatment initiation during the 14-day trial period. After IPT-weighting, the groups were well balanced (standardized mean differences <0.01 for nearly all covariates for A1 vs A2).

**Table 1.** Inverse Probability of Treatment Weighted (IPTW) Baseline Characteristics at First Treatment Assignment to A1 or A2 During the 14-Day Trial Period

| Baseline Characteristics | Treatment Strategy |  | SMD |
| --- | --- | --- | --- |
|  | A1 - Atorvastatin | A2 - Other |  |
| ***Cohort (N)*** | 39,948 | 19,182 |  |
| ***Demographics*** | %/mean(SD)median | %/mean(SD)median |  |
| Age (years, continuous) | 76.7 (8.2) 75.0 | 76.8 (8.3) 75.0 | 0.0056 |
| Age (years, categorical) |  |  | 0.0004 |
| 65-74 | 45.9% | 46.2% |  |
| 75-84 | 34.4% | 33.8% |  |
| 85+ | 19.7% | 19.9% |  |
| Female sex | 51.9% | 52.0% | 0.0018 |
| Race/Ethnicity |  |  | 0.0100 |
| White | 83.7% | 83.6% |  |
| Black | 7.1% | 7.2% |  |
| Asian | 2.3% | 1.8% |  |
| Hispanic | 4.6% | 4.9% |  |
| Year of Index Hospitalization |  |  | 0.0285 |
| 2017 | 33.1% | 34.6% |  |
| 2018 | 33.6% | 32.9% |  |
| 2019 | 33.3% | 32.4% |  |
| ***Characteristics at 1st Medicare Enrollment*** |  |  |  |
| Dual Medicare/Medicaid | 17.2% | 17.5% | 0.0091 |
| Disabled | 11.6% | 11.7% | 0.0040 |
| End stage renal disease | 2.0% | 2.0% | 0.0018 |
| ***Healthcare Research Utilization & Costs*** |  |  |  |
| Inpatient admission (yes/no) | 20.5% | 20.6% | 0.0038 |
| # inpatient admissions | 1.3 (0.8) 1.0 | 1.3 (0.8) 1.0 | 0.0056 |
| # inpatient days | 6.6 (6.9) 4.0 | 6.7 (7.2) 4.0 | 0.0092 |
| ED visit (yes/no) | 44.1% | 44.3% | 0.0051 |
| # Emergency department visits | 0.8 (1.3) 0.0 | 0.8 (1.5) 0.0 | 0.0064 |
| # Unique days w/healthcare encounter | 24 (19) 19 | 24 (19) 19 | 0.0043 |
| Total healthcare costs (USD) | 26,407 (24,929) 18,867 | 26,457 (25,298) 18,863 | 0.0020 |
| ***Baseline Comorbidities*** |  |  |  |
| Charlson Comorbidity Index (continuous) | 6.2 (2.9) 6.0 | 6.2 (2.9) 6.0 | 0.0051 |
| Essential hypertension | 83.5% | 83.6% | 0.0033 |
| Hypertension with complications | 42.1% | 42.2% | 0.0027 |
| Acute myocardial infarction | 66.2% | 65.9% | 0.0065 |
| Nonspecific chest pain | 42.4% | 42.5% | 0.0018 |
| Cardiac dysrhythmias | 37.2% | 37.2% | 0.0010 |
| Heart failure | 29.5% | 29.5% | 0.0002 |
| Cerebral infarction | 36.7% | 36.5% | 0.0044 |
| Esophageal disorders | 32.0% | 32.0% | 0.0006 |
| Thyroid disorders | 24.9% | 24.9% | 0.0015 |
| Diabetes mellitus without complication | 26.9% | 26.8% | 0.0022 |
| Diabetes mellitus, Type 2 | 31.1% | 31.0% | 0.0021 |
| Disorders of lipid metabolism | 68.8% | 68.8% | 0.0004 |
| Fluid and electrolyte disorders | 30.5% | 30.6% | 0.0019 |
| Implant, device or graft related encounter | 41.9% | 41.7% | 0.0037 |
| Personal history of nicotine dependence | 32.3% | 32.2% | 0.0023 |
| Personal history of malignant neoplasm | 22.0% | 22.1% | 0.0006 |
| Chronic kidney disease | 24.4% | 24.4% | 0.0000 |
| Osteoarthritis | 28.1% | 28.1% | 0.0006 |
| Musculoskeletal pain, not low back pain | 32.3% | 32.8% | 0.0101 |
| Spondylopathies/spondyloarthropathy | 23.1% | 23.2% | 0.0012 |
| Malaise and fatigue | 24.9% | 24.9% | 0.0002 |
| Nervous system signs and symptoms | 40.7% | 41.2% | 0.0086 |
| Circulatory signs and symptoms | 22.8% | 22.7% | 0.0014 |
| Respiratory signs and symptoms | 34.5% | 34.7% | 0.0037 |
| Abnormal findings without diagnosis | 46.0% | 46.1% | 0.0020 |
| ***Baseline Pharmacotherapy*** |  |  |  |
| Anti-infective Agents | 45.2% | 45.6% | 0.0082 |
| Autonomic Drugs | 45.7% | 45.9% | 0.0035 |
| Selective Beta Adrenergic Blocking Agent | 25.2% | 25.2% | 0.0005 |
| Class II Antiarrhythmics | 23.4% | 23.3% | 0.0018 |
| Vasodilating Agents, Miscellaneous | 21.4% | 21.4% | 0.0006 |
| Beta Adrenergic Blocking Agents | 32.8% | 32.9% | 0.0002 |
| Angiotensin Converting Enzyme Inhibitors | 21.7% | 21.7% | 0.0001 |
| Central Nervous System Agents | 54.5% | 54.8% | 0.0073 |
| Eye, Ear, Nose, and Throat Preparations | 45.6% | 45.9% | 0.0053 |
| Antibacterials | 26.7% | 26.9% | 0.0048 |
| Corticosteroids | 23.6% | 23.7% | 0.0019 |
| Gastrointestinal Drugs | 42.4% | 42.7% | 0.0066 |
| Proton Pump Inhibitors | 24.1% | 24.3% | 0.0057 |
| Hormones and Synthetic Substitutes | 50.8% | 51.0% | 0.0049 |
| Adrenals | 27.8% | 28.0% | 0.0046 |
| ***Baseline Procedures*** |  |  |  |
| Percutaneous coronary interventions (PCI) | 38.2% | 37.8% | 0.0077 |
| Cardiac and coronary fluoroscopy | 52.5% | 52.3% | 0.0049 |
| Measurement during cardiac catheterization | 48.1% | 47.9% | 0.0030 |
| Routine chest X-ray | 21.1% | 21.2% | 0.0032 |
| Electrocardiogram | 38.7% | 38.9% | 0.0031 |

Abbreviations: SMD = absolute value of the standardized mean difference (A1 vs A2); ED = emergency department;
All baseline comorbidities, healthcare utilization, pharmacotherapy, and procedure variables were ascertained in the 12 months prior to or on the time zero event date (baseline period). The Treatment Strategy columns indicate new initiation of atorvastatin in addition to other prevalent or incident medications (A1) versus new initiation of a medication other than atorvastatin in addition to other prevalent or incident medications (A2) within 14 days post-discharge trial period.

### Outcomes and Sequential Trials

In the overall cohort, we analyzed 295 of the 552 CCSR-derived outcomes after excluding those with fewer than 20 events among patients assigned to treatment strategies A1 or A2 across all sequential trials, those representing sequela or follow-up events of a previously diagnosed condition, and the indication-related outcome (CCSR END010: Disorders of Lipid Metabolism). The number of sequential trials per outcome ranged from 2 to 12 (mean 11, SD 1.4; median 11). Outcomes with higher baseline prevalence or earlier occurrence following the time zero event permitted fewer sequential trials, because a larger proportion of patients were excluded as prevalent cases and because the minimum cell-size requirement (40 patients assigned to each treatment strategy within any given trial) was more often unmet.

### ADE Signal Detection Results

In weighted Fine-Gray models treating death as a competing risk, most subdistribution hazard ratios comparing atorvastatin initiation (A1) with initiation of a different medication (A2; reference) across 295 outcomes were null after the multiplicity correction, with the vast majority showing no association. Five outcomes met the prespecified, primary ADE signal detection criteria (q ≤ 0.10 and sHR ≥ 1.20), indicating a higher relative hazard among patients who initiated atorvastatin. The sensitivity analysis employing the relaxed FDR threshold (q ≤ 0.2 and sHR ≥ 1.20) detected five additional ADE signals. For all detected ADE signals (primary and sensitivity analysis), **Table 2** includes results from the weighted Fine-Gray subdistribution hazard models; **Table 3** includes results from the descriptive outcomes for trial 0. **Table 4** summarizes the prior knowledge status for each detected ADE signal (Labeled, Published, or Novel), including the three most frequently observed ICD-10-CM codes for each detected ADE signal, and the supporting literature or FDA label reference. Stratum-specific results for the overall cohort, time-stratified analyses, and subgroup analyses are presented in **Figures 2A-2J** and supplemental material **Table S3**. The frequency distribution of ICD-10-CM codes observed during follow-up pertaining to each detected ADE signal are included in supplemental material **Table S4.**

**Table 2.** Detected Adverse Drug Event Signals (Primary and Sensitivity Analysis): Subdistribution Hazard Ratios (sHR) from Weighted Fine-Gray Models Comparing Atorvastatin Initiation (A1) versus Initiation of Other Medications (A2, Reference)

| Detected ADE Signals: Primary Analysis (Primary; $q \leq 0.1$ & $sHR \geq 1.2$) and Sensitivity Analysis (SA; $q \leq 0.2$ & $sHR \geq 1.2$) | | | | | | | | | | | |
| --- | --- | --- | --- | --- | --- | --- | --- | --- | --- | --- | --- |
| Weighted Fine-Gray subdistribution hazard models w/death as a competing risk for exposure contrast: A1 vs A2 (reference) |  |  |  |  |  |  |  |  |  |  |  |
| CCSR | CCSR Description | Stratum | Primary | SA | sHR | LCL | UCL | SE | Z | P | P-BH |
| BLD004 | Acute posthemorrhagic anemia | Age 75-84 years | - | Yes | 1.43 | 1.14 | 1.79 | 0.16 | 3.08 | 0.002 | 0.145 |
| BLD004 | Acute posthemorrhagic anemia | White Race | - | Yes | 1.24 | 1.06 | 1.44 | 0.10 | 2.69 | 0.007 | 0.134 |
| **CIR003** | **Nonrheumatic and unspecified valve disorders** | **Time stratified: 121-182 days** | **Yes** | **Yes** | **1.71** | **1.20** | **2.43** | **0.31** | **2.97** | **0.003** | **0.082** |
| CIR021 | Acute hemorrhagic cerebrovascular disease | Overall | - | Yes | 1.44 | 1.09 | 1.89 | 0.20 | 2.56 | 0.011 | 0.194 |
| CIR021 | Acute hemorrhagic cerebrovascular disease | Time stratified: 1-60 days | - | Yes | 1.55 | 1.13 | 2.13 | 0.25 | 2.73 | 0.006 | 0.145 |
| CIR021 | Acute hemorrhagic cerebrovascular disease | White Race | - | Yes | 1.49 | 1.09 | 2.03 | 0.24 | 2.50 | 0.012 | 0.196 |
| CIR035 | Varicose veins of lower extremity | Time stratified: 1-60 days | - | Yes | 1.78 | 1.18 | 2.67 | 0.37 | 2.77 | 0.006 | 0.145 |
| INJ024 | Sprains and strains, initial encounter | Overall | - | Yes | 1.34 | 1.10 | 1.63 | 0.13 | 2.89 | 0.004 | 0.142 |
| **INJ024** | **Sprains and strains, initial encounter** | **Male** | **Yes** | **Yes** | **1.79** | **1.26** | **2.54** | **0.32** | **3.27** | **0.001** | **0.086** |
| SYM012 | Circulatory signs and symptoms | Black Race | - | Yes | 1.51 | 1.16 | 1.96 | 0.20 | 3.09 | 0.002 | 0.167 |
| SYM014 | Skin/Subcutaneous signs and symptoms | Black Race | - | Yes | 1.86 | 1.22 | 2.84 | 0.40 | 2.86 | 0.004 | 0.167 |
| **SYM015** | **General sensation/perception signs and symptoms** | **Overall** | **Yes** | **Yes** | **1.23** | **1.11** | **1.36** | **0.06** | **3.99** | **0.000** | **0.013** |
| SYM015 | General sensation/perception signs and symptoms | Time stratified: 1-60 days | - | Yes | 1.20 | 1.07 | 1.36 | 0.07 | 3.02 | 0.003 | 0.120 |
| SYM015 | General sensation/perception signs and symptoms | Age 85+ years | - | Yes | 1.46 | 1.18 | 1.81 | 0.16 | 3.43 | 0.001 | 0.111 |
| SYM015 | General sensation/perception signs and symptoms | Male | - | Yes | 1.24 | 1.06 | 1.44 | 0.10 | 2.72 | 0.007 | 0.167 |
| **SYM015** | **General sensation/perception signs and symptoms** | **White Race** | **Yes** | **Yes** | **1.20** | **1.08** | **1.34** | **0.07** | **3.35** | **0.001** | **0.060** |
| SYM015 | General sensation/perception signs and symptoms | Black Race | - | Yes | 1.76 | 1.18 | 2.63 | 0.36 | 2.76 | 0.006 | 0.167 |
| **SYM017** | **Abnormal findings without diagnosis** | **Time stratified: 121-182 days** | **Yes** | **Yes** | **1.55** | **1.18** | **2.05** | **0.22** | **3.11** | **0.002** | **0.082** |
| SYM018 | Prediabetes | Time stratified: 1-60 days | - | Yes | 1.64 | 1.11 | 2.40 | 0.32 | 2.51 | 0.012 | 0.185 |
| **SYM018** | **Prediabetes** | **White Race** | **Yes** | **Yes** | **1.71** | **1.24** | **2.36** | **0.28** | **3.29** | **0.001** | **0.060** |

Abbreviations: A1 = Atorvastatin initiation; A2 = Newly initiated a different medication other than atorvastatin; CCSR = Clinical Classifications Software Refined; sHR = Subdistribution hazard ratio; LCL = Lower confidence limit; UCL = Upper confidence limit; SE = Standard error; Z = Wald Z-statistic ( $\beta$ / SE) from the Fine-Gray subdistribution hazard model; P = original p-value from Fine-Gray model; P-BH = Benjamini-Hochberg adjusted p-value; Primary = ADE signal detected using the primary ADE signal detection algorithm ( $q \leq 0.1$ & $sHR \geq 1.2$ ); SA = ADE signal detected in the sensitivity analysis using the ADE signal detection algorithm ( $q \leq 0.2$ & $sHR \geq 1.2$ )

**Table 3.** Detected Adverse Drug Event Signals (Primary and Sensitivity Analysis): Descriptive Outcomes from Trial 0 for Atorvastatin Initiation (A1) and Initiation of Other Medications (A2, Reference)

| ADE Signals Detected: Primary Analysis (Primary; $q \leq 0.1$ & $sHR \geq 1.2$) and Sensitivity Analysis (SA; $q \leq 0.2$ & $sHR \geq 1.2$) | | | | | | | | | | | | | |
| --- | --- | --- | --- | --- | --- | --- | --- | --- | --- | --- | --- | --- | --- |
| Descriptive Outcomes from Trial 0 |  |  |  |  |  |  |  |  |  |  |  |  |  |
| CCSR | CCSR Description | Stratum | Primary | SA | N | A | B | C | D | CI - A2 | CI - A1 | RD | NNH |
| BLD004 | Acute posthemorrhagic anemia | Age 75-84 years | - | Yes | 14436 | 3221 | 10672 | 89 | 454 | 0.027 | 0.041 | 0.014 | 72 |
| BLD004 | Acute posthemorrhagic anemia | White Race | - | Yes | 35759 | 7858 | 26661 | 222 | 1018 | 0.027 | 0.037 | 0.009 | 107 |
| **CIR003** | **Nonrheumatic and unspecified valve disorders** | **Time stratified: 121-182 days** | **Yes** | **Yes** | **16371** | **1634** | **14330** | **28** | **379** | **0.017** | **0.026** | **0.009** | **112** |
| CIR021 | Acute hemorrhagic cerebrovascular disease | Overall | - | Yes | 44400 | 9911 | 34057 | 54 | 378 | 0.005 | 0.011 | 0.006 | 180 |
| CIR021 | Acute hemorrhagic cerebrovascular disease | Time stratified: 1-60 days | - | Yes | 44400 | 9924 | 34176 | 41 | 259 | 0.004 | 0.008 | 0.003 | 294 |
| CIR021 | Acute hemorrhagic cerebrovascular disease | White Race | - | Yes | 37550 | 8536 | 28673 | 45 | 296 | 0.005 | 0.010 | 0.005 | 201 |
| CIR035 | Varicose veins of lower extremity | Time stratified: 1-60 days | - | Yes | 44516 | 9958 | 34373 | 27 | 158 | 0.003 | 0.005 | 0.002 | 534 |
| INJ024 | Sprains and strains, initial encounter | Overall | - | Yes | 43772 | 9665 | 33286 | 112 | 709 | 0.011 | 0.021 | 0.009 | 106 |
| **INJ024** | **Sprains and strains, initial encounter** | **Male** | **Yes** | **Yes** | **21458** | **4443** | **16657** | **35** | **323** | **0.008** | **0.019** | **0.011** | **89** |
| SYM012 | Circulatory signs and symptoms | Black Race | - | Yes | 2281 | 432 | 1415 | 66 | 368 | 0.133 | 0.206 | 0.074 | 14 |
| SYM014 | Skin/Subcutaneous signs and symptoms | Black Race | - | Yes | 2664 | 581 | 1888 | 24 | 171 | 0.040 | 0.083 | 0.043 | 23 |
| **SYM015** | **General sensation/perception signs and symptoms** | **Overall** | **Yes** | **Yes** | **40054** | **8394** | **28368** | **526** | **2766** | **0.059** | **0.089** | **0.030** | **33** |
| SYM015 | General sensation/perception signs and symptoms | Time stratified: 1-60 days | - | Yes | 40054 | 8531 | 29393 | 389 | 1741 | 0.044 | 0.056 | 0.012 | 81 |
| SYM015 | General sensation/perception signs and symptoms | Age 85+ years | - | Yes | 7464 | 2019 | 4841 | 118 | 486 | 0.055 | 0.091 | 0.036 | 28 |
| SYM015 | General sensation/perception signs and symptoms | Male | - | Yes | 19985 | 3918 | 14518 | 237 | 1312 | 0.057 | 0.083 | 0.026 | 39 |
| **SYM015** | **General sensation/perception signs and symptoms** | **White Race** | **Yes** | **Yes** | **33902** | **7244** | **23888** | **456** | **2314** | **0.059** | **0.088** | **0.029** | **34** |
| SYM015 | General sensation/perception signs and symptoms | Black Race | - | Yes | 2568 | 549 | 1822 | 32 | 165 | 0.055 | 0.083 | 0.028 | 36 |
| **SYM017** | **Abnormal findings without diagnosis** | **Time stratified: 121-182 days** | **Yes** | **Yes** | **8269** | **820** | **6690** | **65** | **694** | **0.073** | **0.094** | **0.021** | **49** |
| SYM018 | Prediabetes | Time stratified: 1-60 days | - | Yes | 43707 | 9889 | 33487 | 34 | 297 | 0.003 | 0.009 | 0.005 | 186 |
| **SYM018** | **Prediabetes** | **White Race** | **Yes** | **Yes** | **36982** | **8498** | **28052** | **43** | **389** | **0.005** | **0.014** | **0.009** | **116** |

Abbreviations: A1 = Atorvastatin initiation; A2 = Newly initiated a different medication other than atorvastatin; N = Total number of patients in the analytic stratum; A = Number of A2 patients without the event; B = Number of A1 patients without the event; C = Number of A2 patients with the event; D = Number of A1 patients with the event; CI-A1 = Cumulative incidence (risk) in the A1 (atorvastatin) group; CI-A2 = Cumulative incidence (risk) in the A2 (comparator) group; RD = Risk difference; NNH = Number needed to harm; Primary = ADE signal detected using the primary ADE signal detection algorithm ( $q \leq 0.1$ & $sHR \geq 1.2$ ); SA = ADE signal detected in the sensitivity analysis using the ADE signal detection algorithm ( $q \leq 0.2$ & $sHR \geq 1.2$ )

**Table 4.** Characterization of Detected ADE Signals: Prior Knowledge Status, Dominant ICD-10-CM Codes, and Supporting Literature

| CCSR | CCSR Description | Prior Knowledge | Top 3 ICD-10-CM Codes | Reference |
| --- | --- | --- | --- | --- |
| BLD004 | Acute posthemorrhagic anemia | P | D62 Acute posthemorrhagic anemia (100%) | Amarenco P et al. (SPARCL Investigators) 2006<br>Martinez AI et al. 2019 |
| CIR003 | Nonrheumatic and unspecified valve disorders | P | I340 Nonrheumatic mitral (valve) insufficiency (45.99%)<br>I350 Nonrheumatic aortic (valve) insufficiency (20.71%)<br>I351 Nonrheumatic aortic (valve) stenosis (9.18%) | Myasoedova VA et al. 2025 |
| CIR021 | Acute hemorrhagic cerebrovascular disease | L, P | I619 Nontraumatic intracerebral hemorrhage, unspecified (24.41%)<br>I609 Nontraumatic subarachnoid hemorrhage, unspecified (14.8%)<br>I6000 Nontraumatic subarachnoid hemorrhage, unspecified (14.1%) | Amarenco P et al. (SPARCL Investigators) 2006 |
| CIR035 | Varicose veins of lower extremity | N | I8393 Asymptomatic varicose veins of bilateral lower extremity (13.35%)<br>I8399 Varicose veins of bilateral lower extremities (11.95%)<br>I8390 Varicose veins of unspecified lower extremity (11.95%) | No original research or mention in the atorvastatin prescribing information |
| INJ024 | Sprains and strains, initial encounter | L, P | S161XXA Strain of muscle, fascia and tendon at neck level (10.01%)<br>S134XXA Sprain of ligaments of cervical spine, initial encounter (7.01%)<br>S133XXA Sprain of ligaments of lumbar spine, initial encounter (6.58%) | Mansi I et al. 2013<br>Eliasson P et al. 2019<br>Cholesterol Treatment Trialists' Collaboration 2022<br>Zheng H et al. 2024 |
| SYM012 | Circulatory signs and symptoms | N | R001 Bradycardia, unspecified (29.52%)<br>R002 Palpitations (18.34%)<br>R00989 Other specified symptoms/signs involving circulatory system (18.13%) | No original research or mention in the atorvastatin prescribing information |
| SYM014 | Skin/Subcutaneous signs and symptoms | L | R21 Rash and other nonspecific skin eruption (18.56%)<br>R200 Anesthesia of skin (16.15%)<br>R202 Paresthesia of skin (16.41%) | Atorvastatin prescribing information |
| SYM015 | General sensation/perception signs & symptoms | L | R42 Dizziness and giddiness (95.78%)<br>R43 Hallucinations, unspecified (1.38%)<br>R440 Visual hallucinations (1.38%) | Atorvastatin prescribing information |
| SYM017 | Abnormal findings without diagnosis | L, P | R931 Abnormal electrocardiogram [ECG] [EKG] (22.13%)<br>R938 Other nonspecific abnormal finding of blood field (14.73%)<br>R7989 Other specified abnormal findings of blood chemistry (8.58%) | Amarenco P et al. (SPARCL Investigators) 2006<br>Chandok T et al. 2023 |
| SYM018 | Prediabetes | L, P | R7303 Prediabetes (100%) | Amarenco P et al. (SPARCL Investigators) 2006<br>Waters DD et al. 2011<br>Cholesterol Treatment Trialists' Collaboration 2024 |

**Abbreviations:** L = Explicitly listed in the current atorvastatin (Lipitor) prescribing information (FDA label); P = Previously reported in original research, observational cohorts, or clinical trials; N = Novel or hypothesis-generating signal with limited or no dedicated prior reports. Multiple designations indicate overlapping evidence (e.g., L, P).
**Notes:** The “Top 3 ICD-10-CM Codes” column shows the most prevalent, observed codes within each CCSR category during follow-up. SPARCL = Stroke Prevention by Aggressive Reduction in Cholesterol Levels trial. FDA label references are from the official DailyMed or accessdata.fda.gov versions (2022–2024). All citations are primary sources or the FDA label as noted.

**Figure 1.**
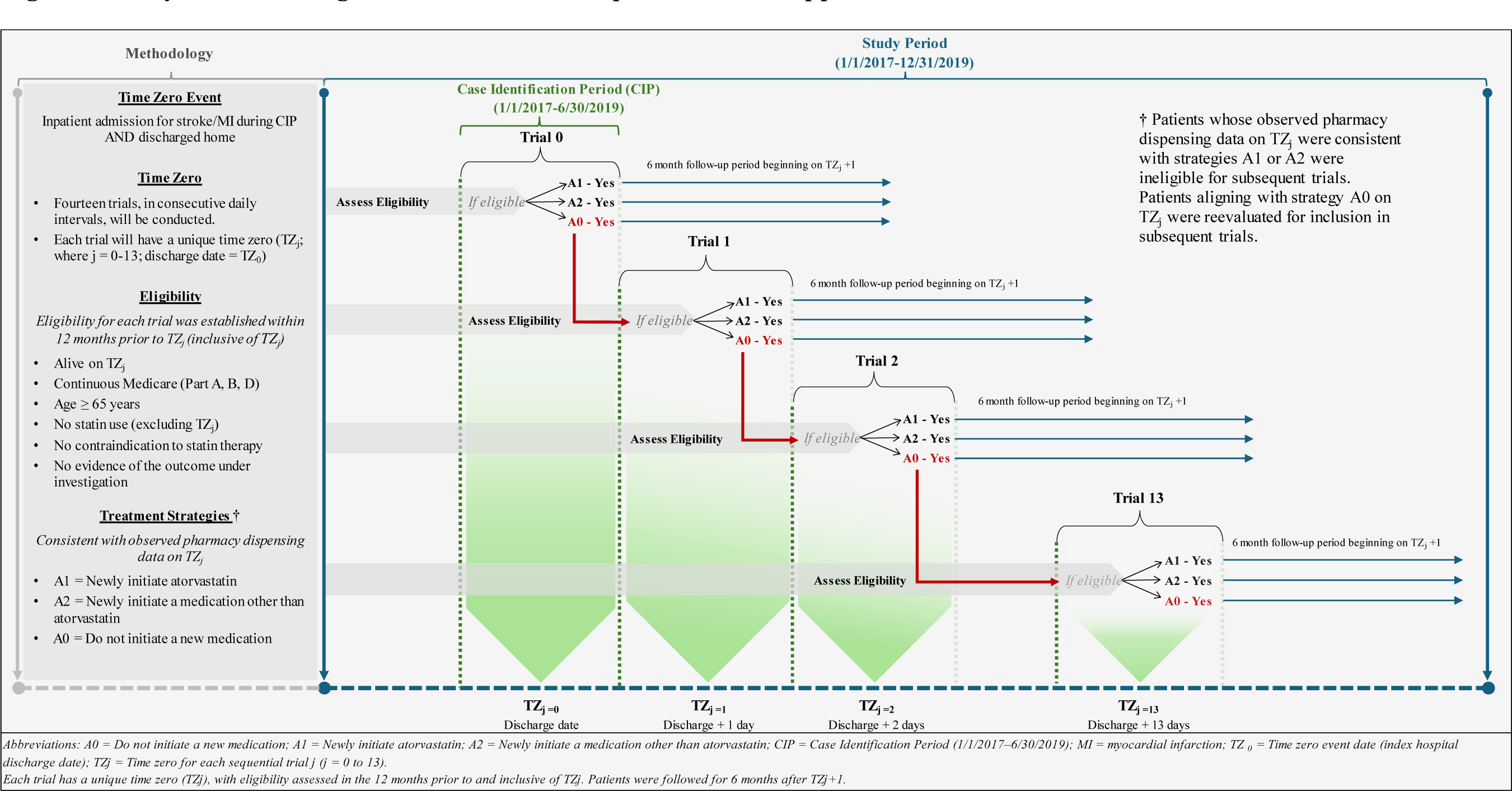
Study Schema: Target Trial Emulation Sequential Trials Approach.

**Figure 2A.**
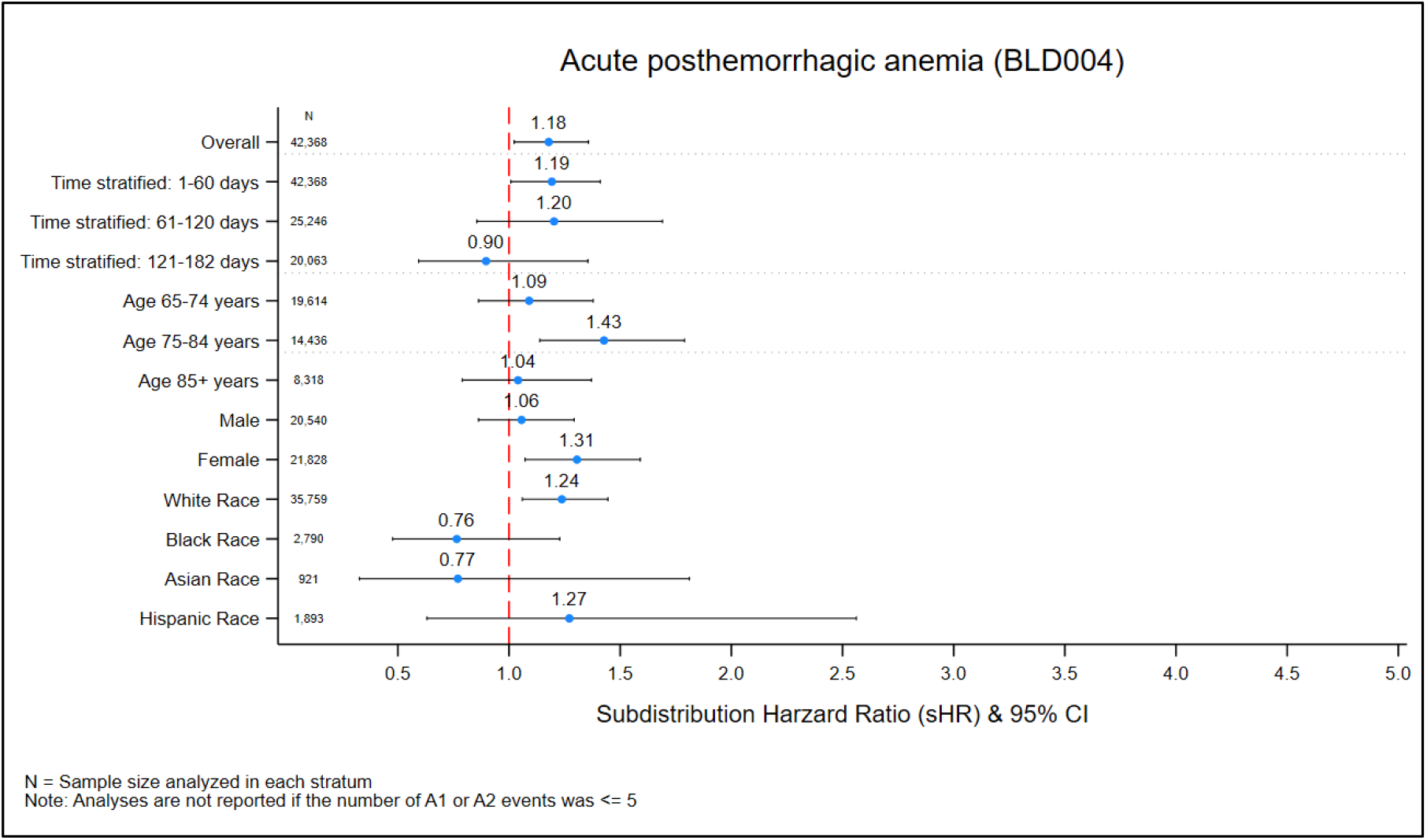
Subdistribution Hazard Ratios for the Detected ADE Signal pertaining to: Acute posthemorrhagic anemia (BLD004)

**Figure 2B.**
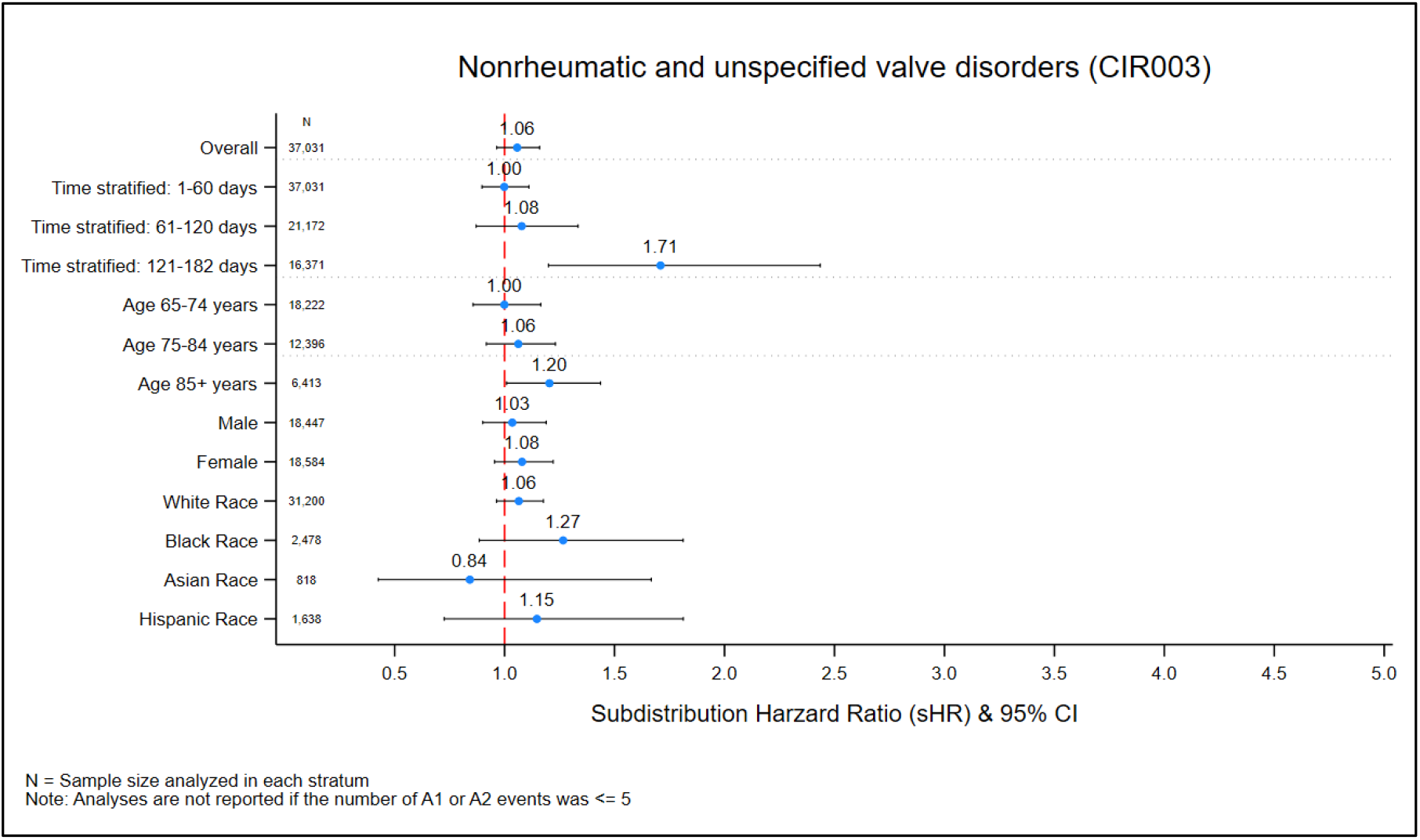
Subdistribution Hazard Ratios for the Detected ADE Signal pertaining to: Nonrheumatic and unspecified valve disorders (CIR003)

**Figure 2C.**
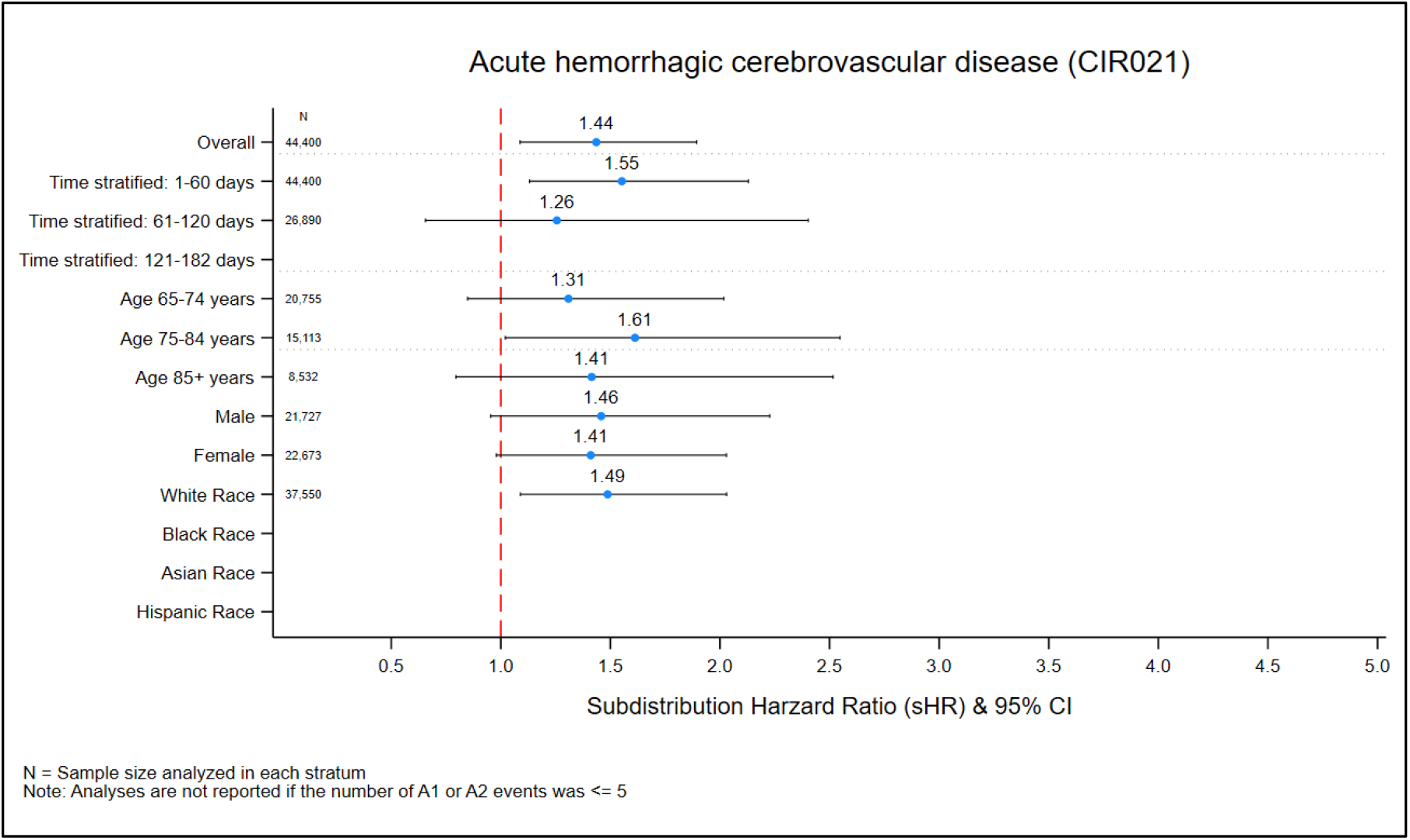
Subdistribution Hazard Ratios for the Detected ADE Signal pertaining to: Acute hemorrhagic cerebrovascular disease (CIR021)

**Figure 2D.**
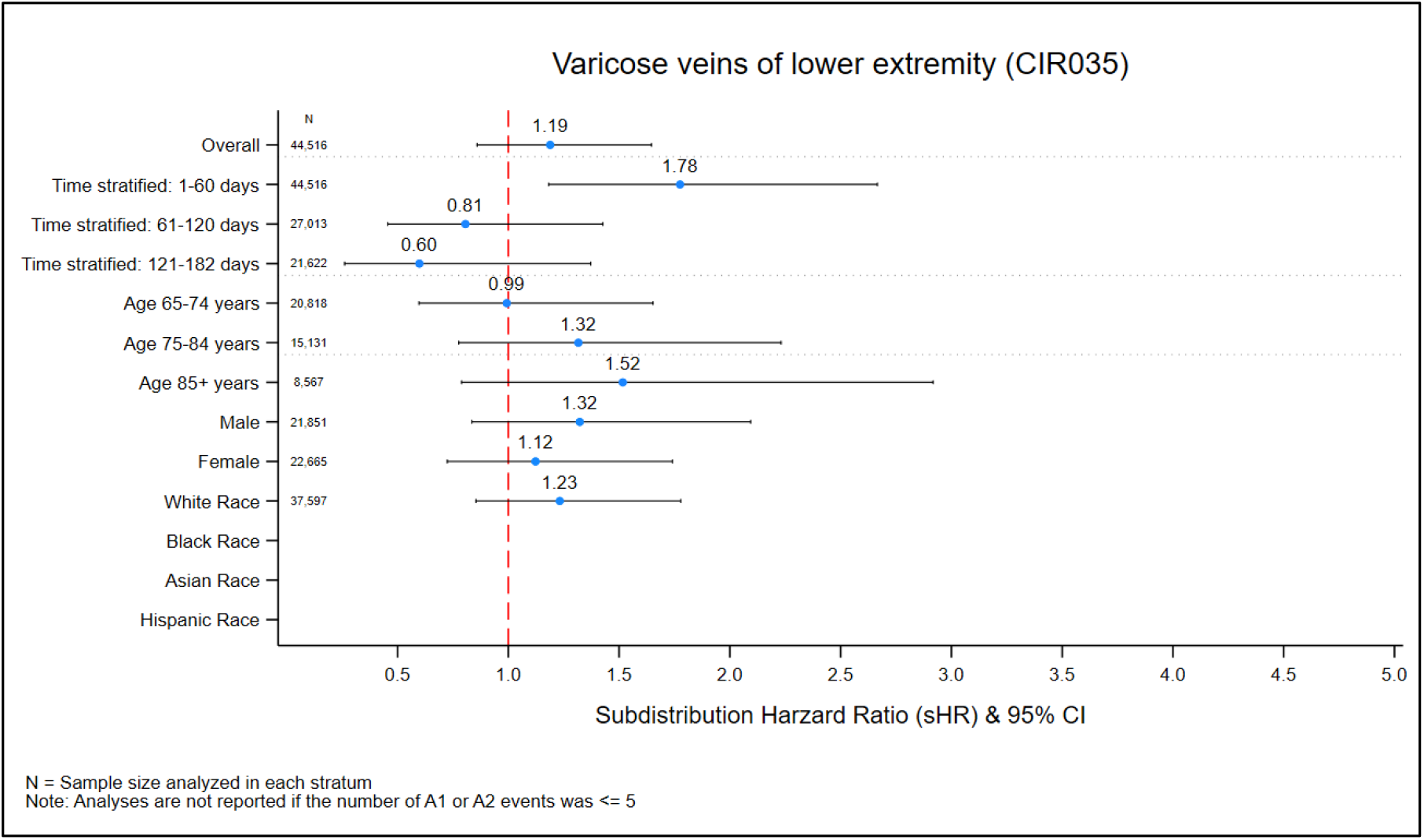
Subdistribution Hazard Ratios for the Detected ADE Signal pertaining to: Varicose veins of lower extremity (CIR035)

**Figure 2E.**
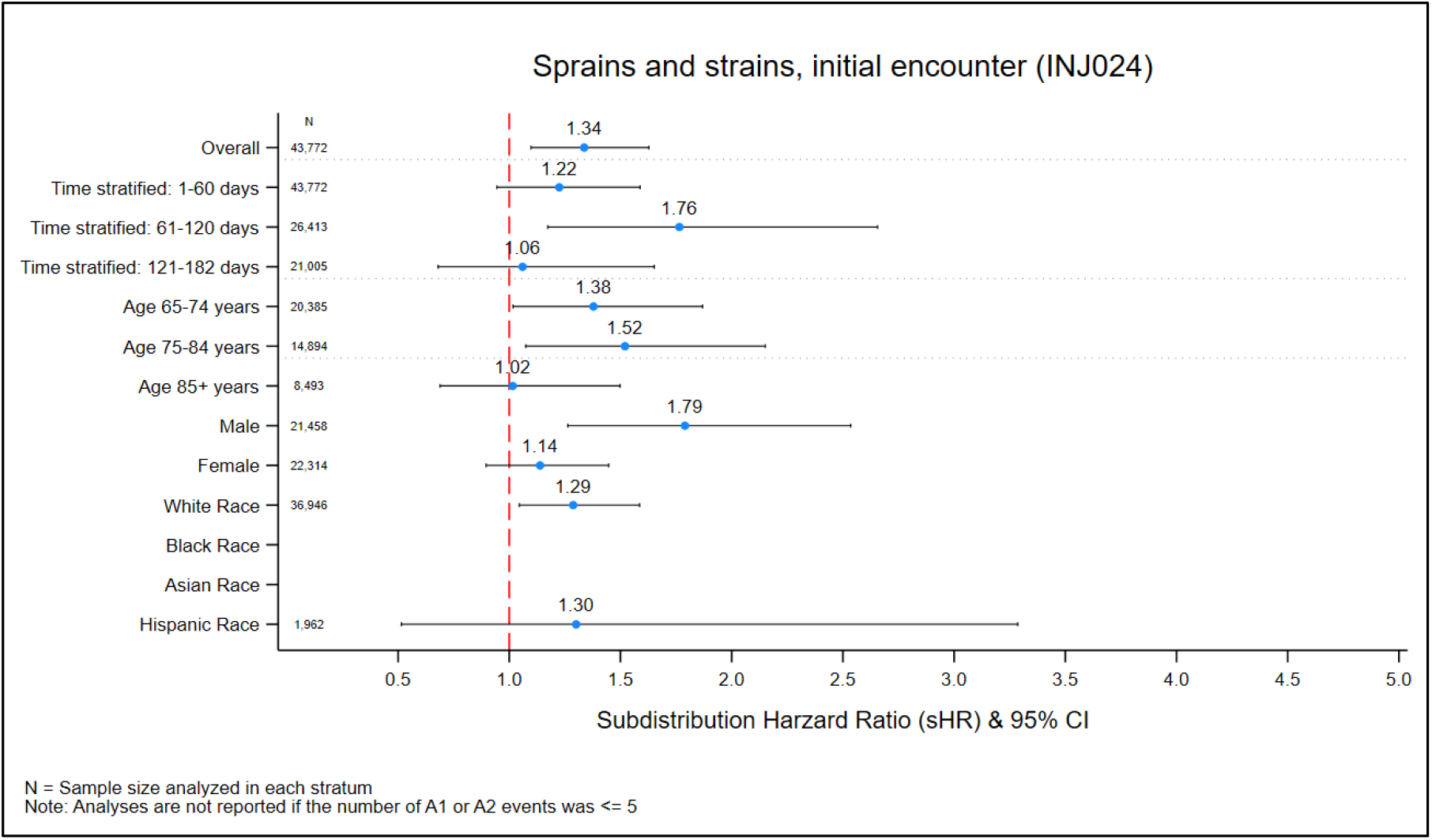
Subdistribution Hazard Ratios for the Detected ADE Signal pertaining to: Sprains and strains, initial encounter (INJ024)

**Figure 2F.**
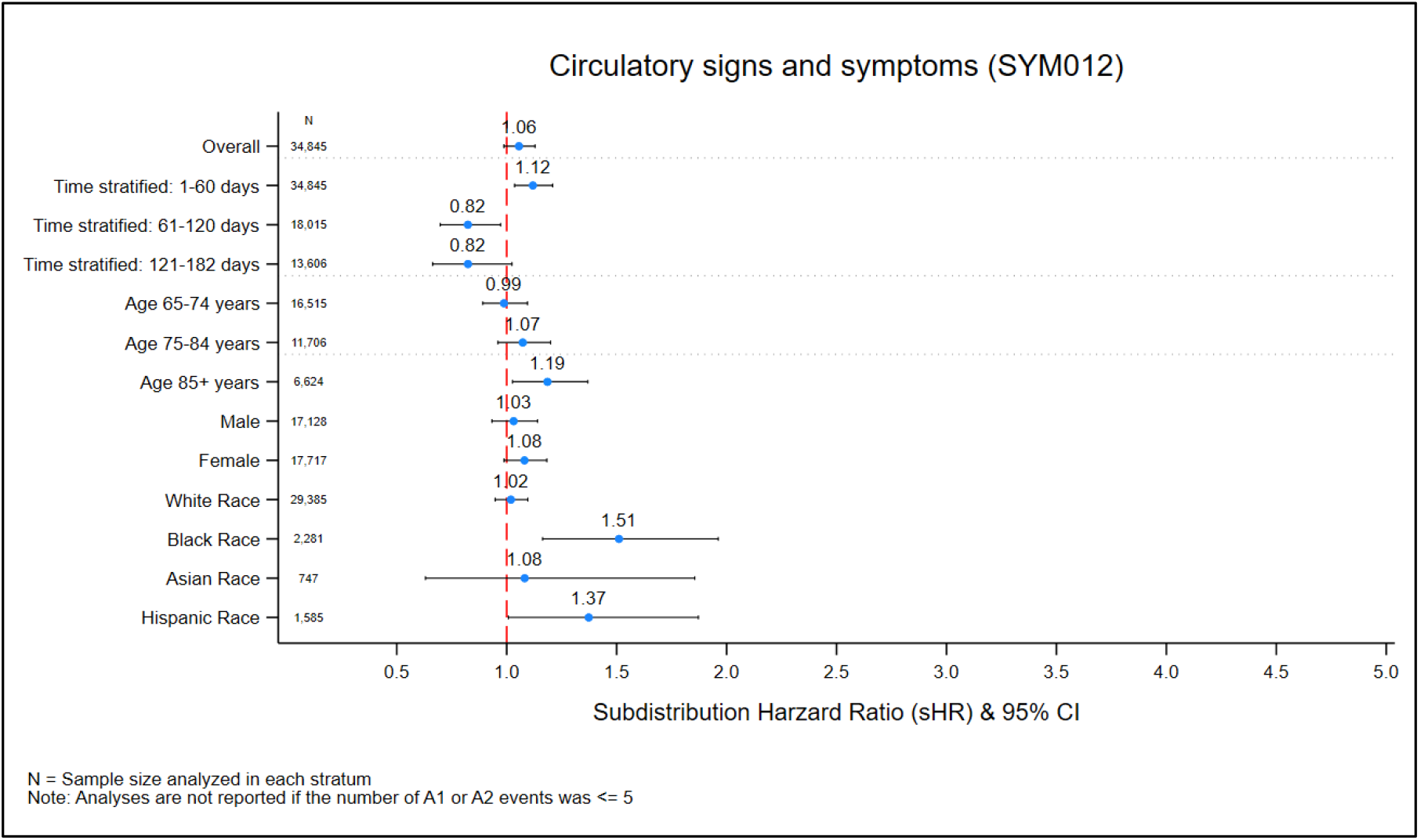
Subdistribution Hazard Ratios for the Detected ADE Signal pertaining to: Circulatory signs and symptoms (SYM012)

**Figure 2G.**
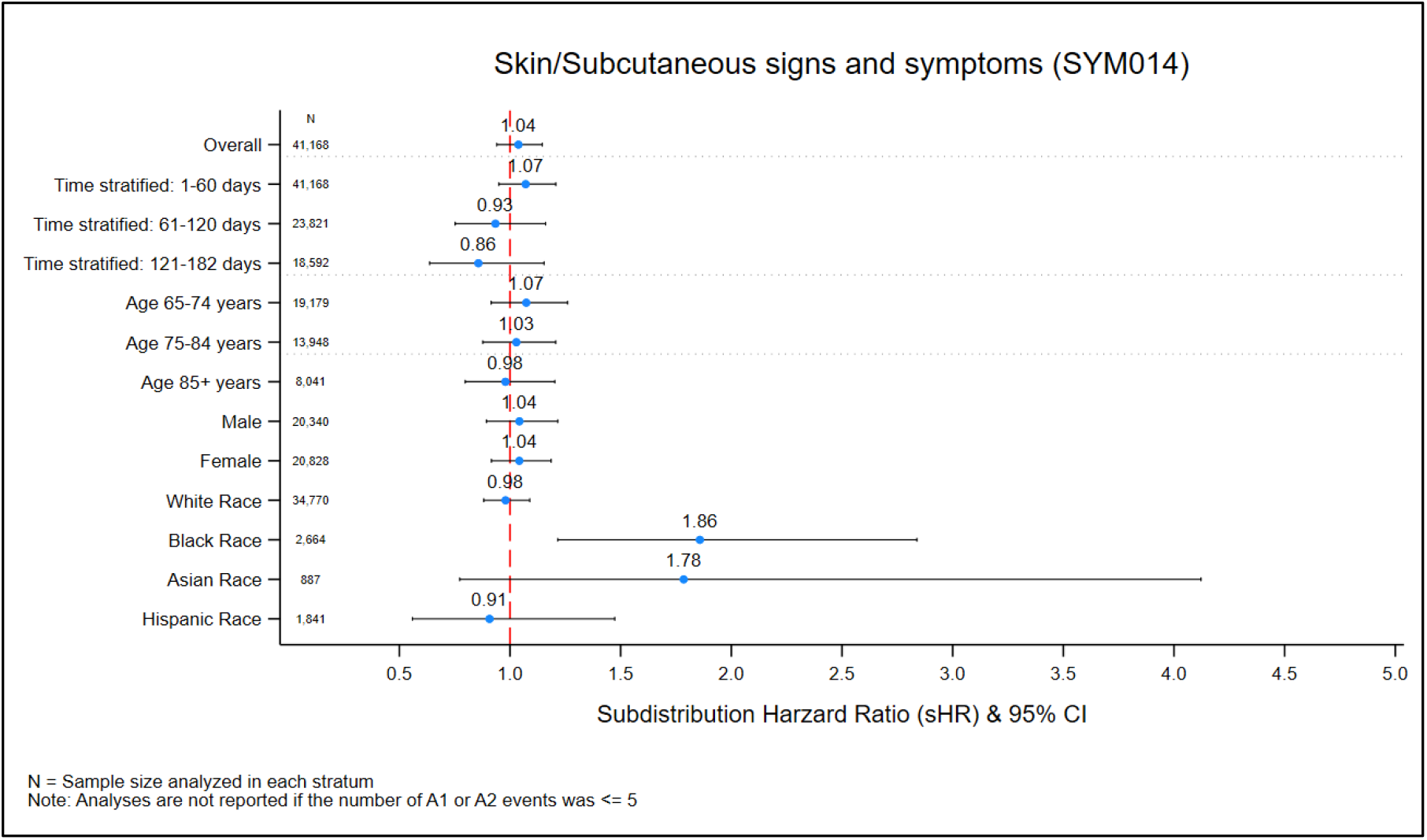
Subdistribution Hazard Ratios for the Detected ADE Signal pertaining to: Skin/Subcutaneous signs and symptoms (SYM014)

**Figure 2H.**
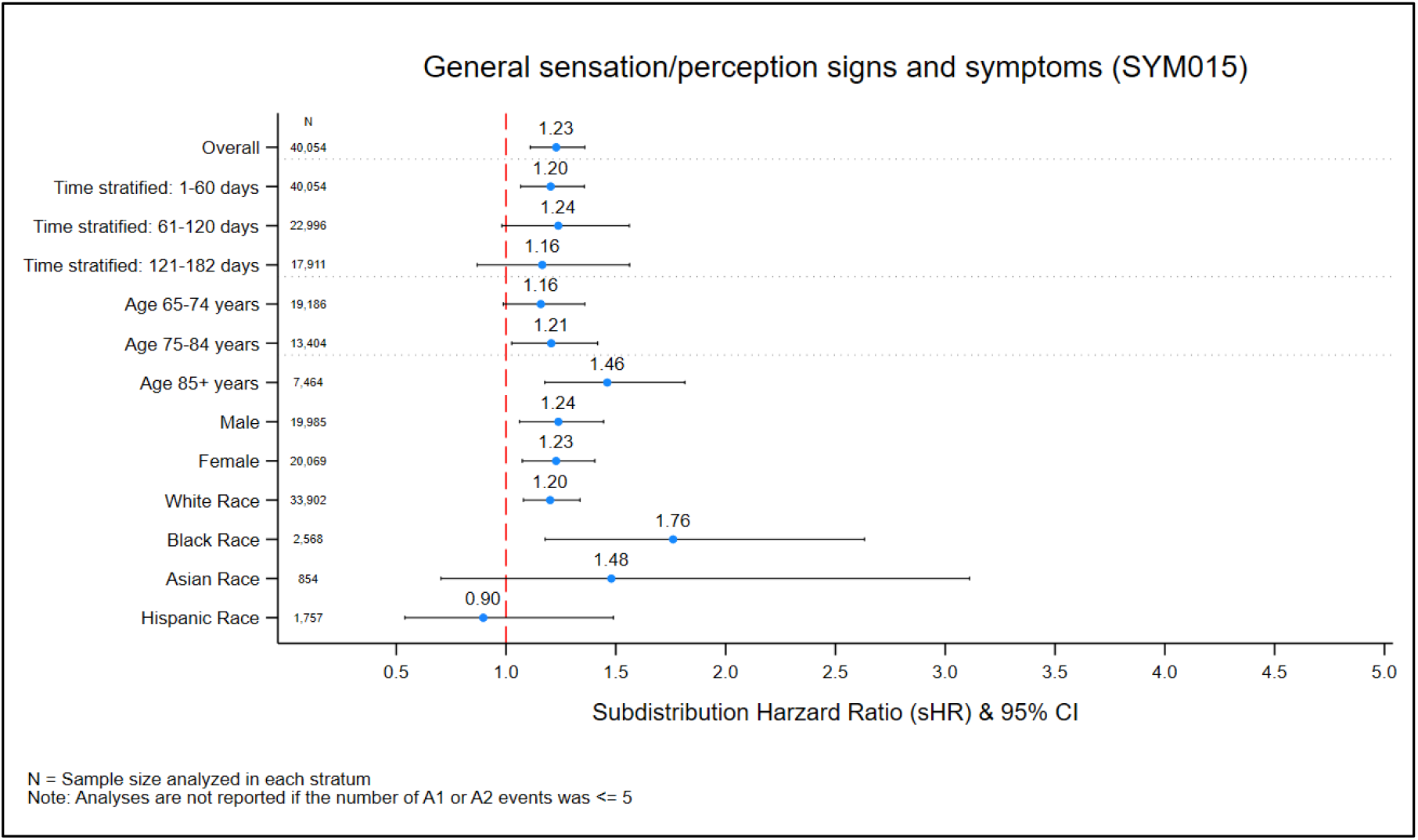
Subdistribution Hazard Ratios for the Detected ADE Signal pertaining to: General sensation/perception signs and symptoms (SYM015)

**Figure 2I.**
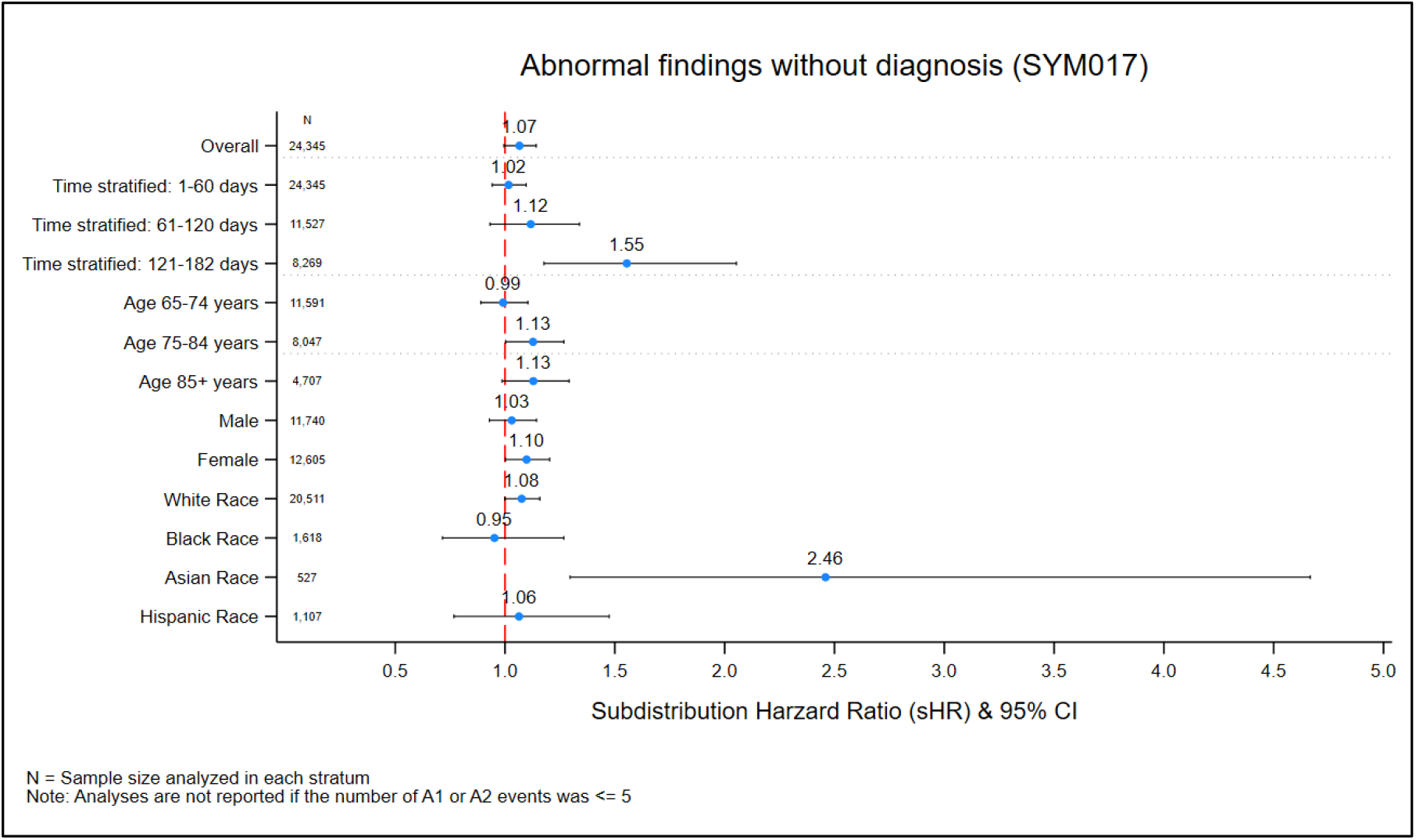
Subdistribution Hazard Ratios for the Detected ADE Signal pertaining to: Abnormal findings without diagnosis (SYM017)

**Figure 2J.**
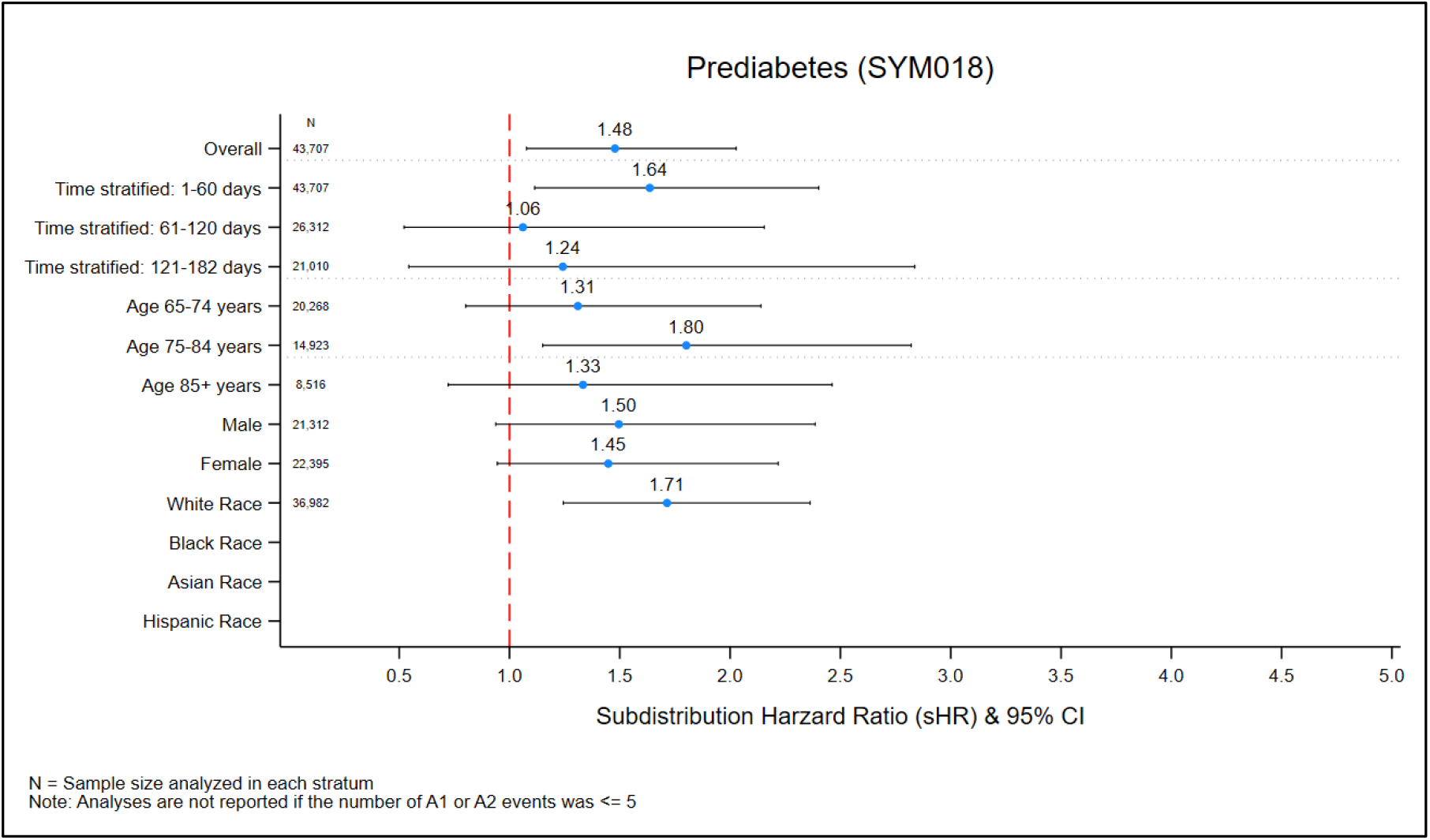
Subdistribution Hazard Ratios for the Detected ADE Signal pertaining to: Prediabetes (SYM018)

Among the five signals meeting the primary signal detection criteria, nonrheumatic and unspecified valve disorders (CIR003) was associated with a higher hazard in the 121–182 day window (sHR, 1.71; 95% CI, 1.20–2.43). In trial 0, the cumulative incidence reached 2.6% among atorvastatin initiators and 1.7% among those starting an alternative agent (risk difference of 0.009 and number needed to harm of 112). Sprains and strains (INJ024) showed an elevated hazard among men (sHR, 1.79; 95% CI, 1.26–2.54), with cumulative incidences of 1.9% and 0.8% (risk difference, 0.011; NNH, 89). General sensation and perception signs and symptoms (SYM015) had higher hazards both overall (sHR, 1.23; 95% CI, 1.11–1.36) and in White patients (sHR, 1.20; 95% CI, 1.08–1.34); corresponding cumulative incidences were 8.9% versus 5.9% (risk difference, 0.03; NNH, 33). Abnormal findings without a diagnosis (SYM017) exhibited an increased hazard in the later period (sHR, 1.55; 95% CI, 1.18–2.05), with cumulative incidences of 9.4% and 7.3% (risk difference, 0.021; NNH, 49). Prediabetes (SYM018) was more frequent among White patients (sHR, 1.71; 95% CI, 1.24–2.36; cumulative incidence 1.4% versus 0.5%; risk difference 0.009; NNH 116).

In sensitivity analyses that relaxed the FDR threshold to q ≤ 0.20, five additional signals were detected. Acute hemorrhagic cerebrovascular disease (CIR021) exhibited an elevated sHR overall (sHR, 1.44; 95% CI, 1.09–1.89), with cumulative incidences of 1.1% among patients who initiated atorvastatin and 0.5% in the reference group (risk difference, 0.006; number needed to harm, 180). This signal was further detected in White beneficiaries (sHR, 1.40; 95% CI, 1.09– 2.03) and early during follow-up (sHR, 1.55; 95% CI, 1.13–2.13). The remaining signals comprised acute posthemorrhagic anemia (BLD004) among patients aged 75–84 years, an early increase in varicose veins of the lower extremity (CIR035), and a higher relative hazard for both circulatory signs and symptoms (SYM012) and skin and subcutaneous signs and symptoms (SYM014) among Black patients.

## DISCUSSION

This study applied an active, claims-based pharmacovigilance framework using sequential target trial emulation to screen hundreds of potential ADEs following atorvastatin initiation among older Medicare beneficiaries discharged home after acute myocardial or cerebral infarction. Among the 59,130 beneficiaries who initiated atorvastatin or another new medication, inverse probability weighting produced close balance between the two pragmatic treatment strategies (A1 and A2). In weighted Fine-Gray analyses, accounting for death as a competing risk, the great majority of subdistribution hazard ratios were near 1.0, consistent with no material association with atorvastatin initiation. Five outcomes nonetheless met the prespecified, primary ADE signal detection criteria (q ≤ 0.10 and sHR ≥ 1.20): nonrheumatic valve disorders, sprains and strains, general sensation and perception symptoms, abnormal findings without diagnosis, and prediabetes. Five additional ADE signals were detected in the sensitivity analysis employing the relaxed FDR threshold (q ≤ 0.20 and sHR ≥ 1.20): acute posthemorrhagic anemia, acute hemorrhagic cerebrovascular disease, varicose veins of lower extremity, circulatory signs and symptoms, and skin/subcutaneous signs and symptoms.

The ten detected ADE signals occupied a wide gradient of causal interpretability that closely tracked existing knowledge about statin-associated ADEs (**Table 4**). Prediabetes (SYM018) and sprains and strains (INJ024) anchored one end as well-characterized class effects: the former served as a positive control whose elevation among White patients matched meta-analytic evidence^32^ and the increased incidence of new-onset diabetes observed in the SPARCL trial with high-dose atorvastatin;^33,45^ the latter aligned with documented musculoskeletal effects on muscle strength, balance, and connective tissue, including the male predominance noted in prior cohort and Mendelian randomization studies.^39,46–48^ Acute hemorrhagic cerebrovascular disease (CIR021), detected only in sensitivity analyses, occupied an equally informative position: it is explicitly listed in the atorvastatin prescribing information and consistent with the increased risk of hemorrhagic stroke observed in the SPARCL trial, an association that persisted in exploratory analyses excluding patients with a prior hemorrhagic event.^33^ The detected ADE signal pertaining to general sensation and perception symptoms (SYM015) was driven almost entirely by “dizziness and giddiness”, a labeled adverse reaction whose central mechanism is biologically plausible for lipophilic statins.

At the opposite pole lay the heterogeneous category of abnormal findings without diagnosis (SYM017), which the sequential trial emulation productively surfaces as several hypotheses warranting targeted confirmation rather than purporting to adjudicate definitively; although glucose- and transaminase-related codes overlap mechanistically with known glycemic and hepatobiliary effects.^49^ The abnormal-electrocardiogram plurality for this outcome is harder to attribute; it could reflect downstream cardiac consequences of statin-related myositis/rhabdomyolysis^50^ or, as plausibly, routine cardiac surveillance in a recently infarcted cohort. The breadth of this outcome category is exactly why it resists a single interpretation and warrants confirmation using more homogeneous outcome definitions to rule out channeling or surveillance bias.

Nonrheumatic valve disorders (CIR003) and the remaining signals fall in intermediate positions: the former showed elevation in late follow-up, with supportive in vitro and observational data on valve calcification and PCSK9 secretion,^51^ yet a post-hoc analysis of randomized atorvastatin trials found no effect on incident aortic valve stenosis and the dominant mitral code lacks explanation.^52^ Acute posthemorrhagic anemia (BLD004) similarly occupied an intermediate position on the gradient, potentially reflecting a downstream consequence of either intracranial hemorrhage (as observed in the SPARCL trial) or extracranial bleeding events such as gastrointestinal hemorrhage. Consistent with this interpretation, a large retrospective cohort study found statin users to have an elevated risk of gastrointestinal hemorrhage, particularly in the first year of treatment,^53^ which may culminate in posthemorrhagic anemia in the vulnerable post-infarction population often receiving concomitant antithrombotic therapy. Each of these associations invites careful scrutiny and additional research in light of unmeasured or residual confounding, competing risks, and the inherent limits of claims-based outcome ascertainment, underscoring both the framework’s capacity to recover known ADEs as validation of its performance and its ability to flag signals that remain incompletely understood [e.g., varicose veins (CIR035) and circulatory symptoms (SYM012)].

Strengths of this study stem from its use of a well-defined, population-based cohort of statin-naïve new users drawn from Medicare beneficiaries with comprehensive longitudinal data on enrollment, comorbidities, medications, healthcare utilization, and diagnosis codes to classify hundreds of potential ADE outcomes. Sequential emulation of target trials, with eligibility and covariates updated at each daily trial origin, mitigated immortal time and channeling bias, produced per-protocol estimates that accounted for informative censoring and competing mortality risk, and anchored every contrast to an empirically defined time zero. Outcomes were ascertained directly from inpatient, emergency department, and outpatient claims, and the pragmatic treatment strategies reflect typical post-discharge polypharmacy, enhancing applicability to routine practice.

Unlike spontaneous reporting systems such as FAERS, which lack denominators needed to accurately estimate cumulative incidence and incidence rates, this active framework using comprehensive longitudinal claims data enabled quantitative evaluation of both known and previously unrecognized ADEs with explicit control of baseline confounding and selection bias (e.g., immortal time bias, and censoring bias). By emulating up to 14 sequential trials for each outcome, we obtained per-protocol effect estimates and confidence intervals for hundreds of outcomes without analyst-driven selection, providing a more reliable assessment of medication-related harm than passive surveillance alone. A particular strength of this approach lies in its capacity to recover several well-documented adverse effects of atorvastatin—such as musculoskeletal toxicity, prediabetes, and hemorrhagic stroke—thereby validating the sequential target trial emulation framework against established knowledge while simultaneously flagging signals like nonrheumatic valve disorders that may represent important but incompletely characterized risks and generating credible hypotheses for further causal investigation. All analyses adhered strictly to the prespecified protocol and statistical analysis plan, with no major deviations or post-hoc modifications to the eligibility criteria, treatment definitions, outcome algorithms, signal-detection thresholds, or analytic populations.

Several limitations warrant consideration. The Clinical Classifications Software Refined categories are clinically heterogeneous for some outcomes, most evidently abnormal findings without diagnosis (SYM017). Planned confirmatory research will disaggregate these categories into more homogeneous outcomes, apply a stricter false discovery rate threshold, consider narrower reference groups, and impose more granular exclusion criteria; only sufficiently powered analyses will be considered in the hypothesis-driven confirmatory study. The primary signal detection algorithm (i.e., q ≤ 0.10 and sHR ≥ 1.2) threshold prioritized sensitivity over specificity, an appropriate application for exploratory, high-dimensional ADE screening. However, the absence of detected signals for several labeled statin ADEs—including cognitive impairment, depression, sleep disturbance, and peripheral neuropathy—is consistent with a recent meta-analysis of 19 large statin trials showing no evidence of causal associations,^54^ both validating the present study’s sequential target trial emulation pharmacovigilance framework and reinforcing that atorvastatin does not materially contribute to these adverse events in routine practice among older adults.

Despite extensive adjustment for measured covariates at each trial origin, residual confounding by unmeasured factors such as LDL cholesterol, body mass index, frailty, and lifestyle behaviors cannot be excluded. Although treatment strategy A2 encompassed a heterogeneous set of medications, the distributions of other newly initiated drugs were generally comparable between the A1 and A2 groups. In the A2 group, the five most commonly initiated medications were clopidogrel (38% in A2 vs 46% in A1), nitroglycerin (21% vs 25%), metoprolol tartrate (18% vs 24%), furosemide (17% vs 16%), and lisinopril (17% vs 24%). These patterns suggest that heterogeneity within treatment strategy A2 did not introduce major imbalance in concomitant pharmacotherapy. Finally, the findings were derived from fee-for-service Medicare beneficiaries and may not generalize to Medicare Advantage enrollees or to younger or commercially insured populations.

## CONCLUSION

We emulated sequential pragmatic target trials using Medicare claims data to evaluate the causal effect of atorvastatin initiation, compared with other newly initiated medications, on potential adverse outcomes among statin-naïve older adults following myocardial or cerebral infarction. The detected ADE signals that emerged fell along a clear gradient of causal plausibility, ranging from well-characterized statin class effects (prediabetes and sprains and strains) and outcomes already noted in product labeling or prior trials (acute hemorrhagic cerebrovascular disease and dizziness) to more heterogeneous and tentative associations (cardiac valve disorders) that resist straightforward interpretation. The sequential target trial emulation framework enabled this assessment by providing per-protocol estimates with explicit control of confounding and by mitigating censoring and immortal time biases—advantages unattainable with passive surveillance systems. This proactive approach has clear strengths but also important limitations, including the clinical heterogeneity of some outcome definitions, the possibility of residual confounding, and restricted generalizability beyond fee-for-service Medicare. The detected ADE signals therefore warrant cautious interpretation and replication in independent data sources with more homogeneous endpoints and stricter exclusion criteria. Overall, this study establishes a scalable, active “Pharmacovigilance 2.0” framework grounded in causal inference principles that recovers known ADEs—thereby validating its performance in a high-risk population—while flagging credible hypotheses for confirmatory research.

## ETHICS STATEMENT

The research program pertaining to this study, titled “Pharmacovigilance 2.0: Elderly, Disabled, and Disadvantaged Populations”, was reviewed and approved by the Institutional Review Board (IRB) at Rutgers University Human Research Protection Program (HRPP) on March 12, 2024. The IRB classified the research as exempt under categories 4(ii) and 4(iii) of the U.S. Code of Federal Regulations, Title 45, Part 46 (45 CFR 46), determining it poses minimal risk. Category 4(ii) applies to research using existing, anonymized data or records where participants cannot be identified, while category 4(iii) covers benign behavioral interventions or interactions with adults where identifiable information is protected, and disclosure poses no significant risk. The principal investigator, Christopher G. Rowan PhD, conducted the study with approval under eIRB protocol #Pro2024000224, supported by the Veritas Research Foundation.

## DATA AVAILABILITY STATEMENT

This study was completed in part by data resources provided by the Institute for Health Data Core at Rutgers University, available at: http://www.ifhcore.rutgers.edu. The data supporting the research initiative, titled *Pharmacovigilance 2.0: Elderly, Disabled, and Disadvantage Populations* and conducted under the principal investigator Christopher Rowan, were obtained from the Centers for Medicare & Medicaid Services (CMS) under Data Use Agreement number RSCH-2024-70249. These data contain protected health information subject to the Privacy Act and HIPAA regulations. Due to privacy, confidentiality, and contractual restrictions imposed by CMS, the datasets are not publicly available and cannot be shared by the authors. Qualified researchers interested in accessing similar data may apply independently through the Research Data Assistance Center (ResDAC) at https://resdac.org/ and execute their own Data Use Agreement with CMS.

## ACKNOWLEDGEMENTS

We express our sincere gratitude to Jesse Berlin, ScD, Dan Horton, MD, MSCE, and Tobi Gerhard, BSPharm, PhD, FISPE, for their invaluable scientific and methodologic support. We are deeply thankful to Mathew Iozzio, Haoqian Chen PhD, and the Center for Pharmacoepidemiology Treatment Science at Rutgers University for their partnership on Pharmacovigilance 2.0. We also appreciate the Center for Medicare and Medicaid Services (CMS) for providing data access and support for this initiative. Heartfelt thanks go to the Veritas Research Foundation Board of Directors - Scott C. Durbin, MPH, Mathew Allard, MBA, Bradley J. Buecker, MFA, Gregory Lobdell, MA, and LeeAnn Hard, CPA—for their steadfast trust and support, and to U.S. Medicare beneficiaries for trusting the Veritas Research Foundation to conduct independent drug safety research on their behalf.

## SUPPLEMENTAL MATERIAL

**Table S1.**
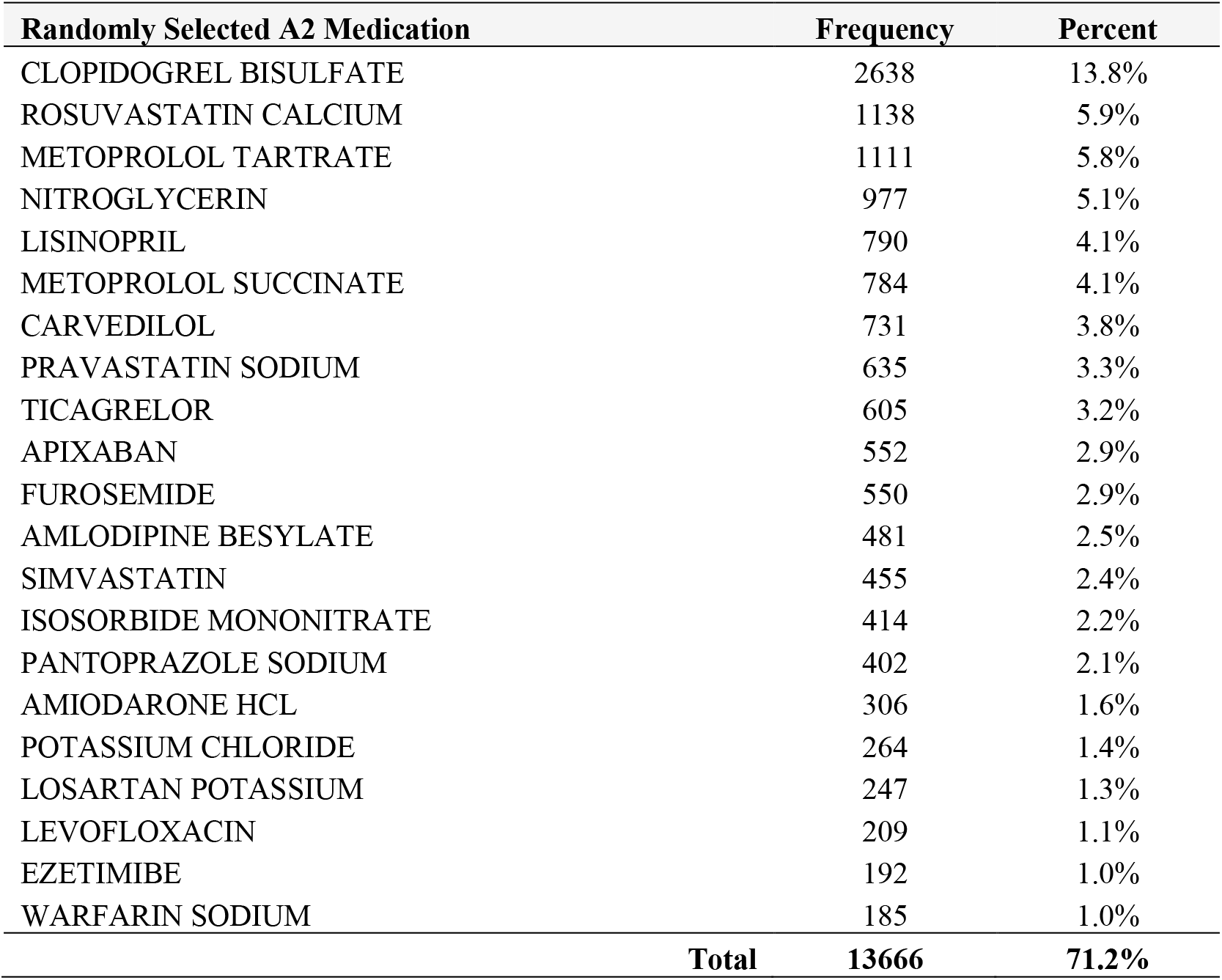
Top 20 Randomly Selected Medications to Classify A2 Discontinuation.

| **Randomly Selected A2 Medication** | **Frequency** | **Percent** |
| --- | --- | --- |
| CLOPIDOGREL BISULFATE | 2638 | 13.8% |
| ROSUVASTATIN CALCIUM | 1138 | 5.9% |
| METOPROLOL TARTRATE | 1111 | 5.8% |
| NITROGLYCERIN | 977 | 5.1% |
| LISINOPRIL | 790 | 4.1% |
| METOPROLOL SUCCINATE | 784 | 4.1% |
| CARVEDILOL | 731 | 3.8% |
| PRAVASTATIN SODIUM | 635 | 3.3% |
| TICAGRELOR | 605 | 3.2% |
| APIXABAN | 552 | 2.9% |
| FUROSEMIDE | 550 | 2.9% |
| AMLODIPINE BESYLATE | 481 | 2.5% |
| SIMVASTATIN | 455 | 2.4% |
| ISOSORBIDE MONONITRATE | 414 | 2.2% |
| PANTOPRAZOLE SODIUM | 402 | 2.1% |
| AMIODARONE HCL | 306 | 1.6% |
| POTASSIUM CHLORIDE | 264 | 1.4% |
| LOSARTAN POTASSIUM | 247 | 1.3% |
| LEVOFLOXACIN | 209 | 1.1% |
| EZETIMIBE | 192 | 1.0% |
| WARFARIN SODIUM | 185 | 1.0% |
| **Total** | **13666** | **71.2%** |

**Table S2.** Reason for Censoring and Follow-up Time by Treatment Strategy before applying inverse probability of censoring weights.

| Reason for End of Follow-up and Distribution of Follow-up Time by Treatment Strategy |  |  |  |  |
| --- | --- | --- | --- | --- |
| Censoring Reason | Treatment Strategy |  |  |  |
|  | A1 | A1 | A2 | A2 |
|  | n | % | n | % |
| Death | 1,578 | 4.0% | 806 | 4.2% |
| Administrative termination (loss of A/B/D coverage) | 464 | 1.2% | 95 | 0.5% |
| Initiated statin (other than atorvastatin) | 2,074 | 5.2% | 6,971 | 36.3% |
| Treatment strategy switch | 5,602 | 14.0% | 2,890 | 15.1% |
| Discontinuation of assigned treatment strategy | 7,484 | 18.7% | 5,167 | 26.9% |
| End of study period before 182 days | 4,632 | 11.6% | 842 | 4.4% |
| Uncensored (completed 182 days of follow-up) | 18,114 | 45.3% | 2411 | 12.6% |
| **Total** | **39,948** | **100.0%** | **19,182** | **100.0%** |

| Time to First Censoring Event | Treatment Strategy |  |
| --- | --- | --- |
|  | A1 | A2 |
| N | 39,948 | 19,182 |
| Mean (SD) | 119 (67) | 52 (64) |
| Median (IQR) | 148 (60–182) | 32 (0–69) |

**Table S3.** Weighted Fine-Gray and Descriptive Analyses for Trial 0 for Detected ADE Signals (Primary Analysis and Sensitivity Analysis)

| Detected ADE Signals: Primary Algorithm ($q \leq 0.1$ & $sHR \geq 1.2$) and Sensitivity Analysis ($q \leq 0.2$ & $sHR \geq 1.2$) | | | | | | | | | | | | | | | | |
| --- | --- | --- | --- | --- | --- | --- | --- | --- | --- | --- | --- | --- | --- | --- | --- | --- |
| CCSR | CCSR Description | Strata | Detected ADE Signal Weighted Fine-Gray |  |  |  |  | Unadjusted Descriptive Analyses for Trial 0 |  |  |  |  |  |  |  |  |
|  |  |  | Primary | SA | sHR | LCL | UCL | N | A | B | C | D | Risk A2 | Risk A1 | RD | NNH |
| BLD004 | Acute posthemorrhagic anemia | Overall | - | - | 1.18 | 1.02 | 1.36 | 42368 | 9146 | 31757 | 261 | 1204 | 0.028 | 0.037 | 0.009 | 114 |
| BLD004 | Acute posthemorrhagic anemia | 1-60 days | - | - | 1.19 | 1.01 | 1.41 | 42368 | 9216 | 32207 | 191 | 754 | 0.020 | 0.023 | 0.003 | 389 |
| BLD004 | Acute posthemorrhagic anemia | 61-120 days | - | - | 1.20 | 0.86 | 1.69 | 25246 | 3363 | 21560 | 39 | 284 | 0.011 | 0.013 | 0.002 | 650 |
| BLD004 | Acute posthemorrhagic anemia | 121-182 days | - | - | 0.90 | 0.59 | 1.35 | 20063 | 2267 | 17599 | 31 | 166 | 0.013 | 0.009 | -0.004 | n/a |
| BLD004 | Acute posthemorrhagic anemia | Age 65-74 years | - | - | 1.09 | 0.86 | 1.38 | 19614 | 3638 | 15357 | 98 | 521 | 0.026 | 0.033 | 0.007 | 152 |
| **BLD004** | **Acute posthemorrhagic anemia** | **Age 75-84 years** | - | **Yes** | **1.43** | **1.14** | **1.79** | **14436** | **3221** | **10672** | **89** | **454** | **0.027** | **0.041** | **0.014** | **72** |
| BLD004 | Acute posthemorrhagic anemia | Age 85+ years | - | - | 1.04 | 0.79 | 1.37 | 8318 | 2287 | 5728 | 74 | 229 | 0.031 | 0.038 | 0.007 | 141 |
| BLD004 | Acute posthemorrhagic anemia | Male | - | - | 1.06 | 0.86 | 1.29 | 20540 | 4136 | 15658 | 126 | 620 | 0.030 | 0.038 | 0.009 | 117 |
| BLD004 | Acute posthemorrhagic anemia | Female | - | - | 1.31 | 1.07 | 1.59 | 21828 | 5010 | 16099 | 135 | 584 | 0.026 | 0.035 | 0.009 | 114 |
| **BLD004** | **Acute posthemorrhagic anemia** | **White Race** | - | **Yes** | **1.24** | **1.06** | **1.44** | **35759** | **7858** | **26661** | **222** | **1018** | **0.027** | **0.037** | **0.009** | **107** |
| BLD004 | Acute posthemorrhagic anemia | Black Race | - | - | 0.76 | 0.48 | 1.23 | 2790 | 601 | 2084 | 24 | 81 | 0.038 | 0.037 | -0.001 | n/a |
| BLD004 | Acute posthemorrhagic anemia | Asian Race | - | - | 0.77 | 0.33 | 1.81 | 921 | 130 | 759 | * | 27 | * | 0.034 | * | * |
| BLD004 | Acute posthemorrhagic anemia | Hispanic Race | - | - | 1.27 | 0.63 | 2.56 | 1893 | 374 | 1459 | * | 54 | * | 0.036 | * | * |
| CIR003 | Nonrheumatic and unspecified valve disorders | Overall | - | - | 1.06 | 0.96 | 1.16 | 37031 | 7037 | 26193 | 640 | 3161 | 0.083 | 0.108 | 0.024 | 41 |
| CIR003 | Nonrheumatic and unspecified valve disorders | 1-60 days | - | - | 1.00 | 0.90 | 1.11 | 37031 | 7168 | 27359 | 509 | 1995 | 0.066 | 0.068 | 0.002 | 602 |
| CIR003 | Nonrheumatic and unspecified valve disorders | 61-120 days | - | - | 1.08 | 0.87 | 1.33 | 21172 | 2431 | 17851 | 103 | 787 | 0.041 | 0.042 | 0.002 | 634 |
| **CIR003** | **Nonrheumatic and unspecified valve disorders** | **121-182 days** | **Yes** | **Yes** | **1.71** | **1.20** | **2.43** | **16371** | **1634** | **14330** | **28** | **379** | **0.017** | **0.026** | **0.009** | **112** |
| CIR003 | Nonrheumatic and unspecified valve disorders | Age 65-74 years | - | - | 1.00 | 0.86 | 1.16 | 18222 | 3114 | 13437 | 227 | 1444 | 0.068 | 0.097 | 0.029 | 34 |
| CIR003 | Nonrheumatic and unspecified valve disorders | Age 75-84 years | - | - | 1.06 | 0.92 | 1.23 | 12396 | 2424 | 8586 | 246 | 1140 | 0.092 | 0.117 | 0.025 | 40 |
| CIR003 | Nonrheumatic and unspecified valve disorders | Age 85+ years | - | - | 1.20 | 1.01 | 1.44 | 6413 | 1499 | 4170 | 167 | 577 | 0.100 | 0.122 | 0.021 | 47 |
| CIR003 | Nonrheumatic and unspecified valve disorders | Male | - | - | 1.03 | 0.90 | 1.19 | 18447 | 3300 | 13338 | 273 | 1536 | 0.076 | 0.103 | 0.027 | 37 |
| CIR003 | Nonrheumatic and unspecified valve disorders | Female | - | - | 1.08 | 0.95 | 1.22 | 18584 | 3737 | 12855 | 367 | 1625 | 0.089 | 0.112 | 0.023 | 44 |
| CIR003 | Nonrheumatic and unspecified valve disorders | White Race | - | - | 1.06 | 0.96 | 1.18 | 31200 | 6018 | 21926 | 552 | 2704 | 0.084 | 0.110 | 0.026 | 39 |
| CIR003 | Nonrheumatic and unspecified valve disorders | Black Race | - | - | 1.27 | 0.88 | 1.81 | 2478 | 498 | 1773 | 39 | 168 | 0.073 | 0.087 | 0.014 | 72 |
| CIR003 | Nonrheumatic and unspecified valve disorders | Asian Race | - | - | 0.84 | 0.43 | 1.67 | 818 | 103 | 631 | 12 | 72 | 0.104 | 0.102 | -0.002 | n/a |
| CIR003 | Nonrheumatic and unspecified valve disorders | Hispanic Race | - | - | 1.15 | 0.73 | 1.81 | 1638 | 287 | 1197 | 20 | 134 | 0.065 | 0.101 | 0.036 | 28 |

**Table S3 Continued: Weighted Fine-Gray and Descriptive Analyses for Trial 0 for Detected ADE Signals (Primary Analysis and Sensitivity Analysis)**
| Detected ADE Signals: Primary Algorithm (q ≤ 0.1 & sHR ≥ 1.2) and Sensitivity Analysis (q ≤ 0.2 & sHR ≥ 1.2) |  |  |  |  |  |  |  |  |  |  |  |  |  |  |  |  |
| --- | --- | --- | --- | --- | --- | --- | --- | --- | --- | --- | --- | --- | --- | --- | --- | --- |
| CCSR | CCSR Description | Strata | Detected ADE Signal Weighted Fine-Gray |  |  |  |  | Unadjusted Descriptive Analyses for Trial 0 |  |  |  |  |  |  |  |  |
|  |  |  | Primary | SA | sHR | LCL | UCL | N | A | B | C | D | Risk A2 | Risk A1 | RD | NNH |
| CIR021 | Acute hemorrhagic cerebrovascular disease | Overall | - | Yes | 1.44 | 1.09 | 1.89 | 44400 | 9911 | 34057 | 54 | 378 | 0.005 | 0.011 | 0.006 | 180 |
| CIR021 | Acute hemorrhagic cerebrovascular disease | 1-60 days | - | Yes | 1.55 | 1.13 | 2.13 | 44400 | 9924 | 34176 | 41 | 259 | 0.004 | 0.008 | 0.003 | 294 |
| CIR021 | Acute hemorrhagic cerebrovascular disease | 61-120 days | - | - | 1.26 | 0.66 | 2.40 | 26890 | 3619 | 23188 | * | 74 | * | 0.003 | * | * |
| CIR021 | Acute hemorrhagic cerebrovascular disease | 121-182 days | - | - | 1.42 | 0.56 | 3.58 | 21539 | 2464 | 19026 | * | 45 | * | 0.002 | * | * |
| CIR021 | Acute hemorrhagic cerebrovascular disease | Age 65-74 years | - | - | 1.31 | 0.85 | 2.02 | 20755 | 3988 | 16579 | 19 | 169 | 0.005 | 0.010 | 0.005 | 187 |
| CIR021 | Acute hemorrhagic cerebrovascular disease | Age 75-84 years | - | - | 1.61 | 1.02 | 2.55 | 15113 | 3488 | 11464 | 20 | 141 | 0.006 | 0.012 | 0.006 | 155 |
| CIR021 | Acute hemorrhagic cerebrovascular disease | Age 85+ years | - | - | 1.41 | 0.80 | 2.52 | 8532 | 2435 | 6014 | 15 | 68 | 0.006 | 0.011 | 0.005 | 198 |
| CIR021 | Acute hemorrhagic cerebrovascular disease | Male | - | - | 1.46 | 0.95 | 2.23 | 21727 | 4532 | 16987 | 23 | 185 | 0.005 | 0.011 | 0.006 | 175 |
| CIR021 | Acute hemorrhagic cerebrovascular disease | Female | - | - | 1.41 | 0.98 | 2.03 | 22673 | 5379 | 17070 | 31 | 193 | 0.006 | 0.011 | 0.005 | 183 |
| CIR021 | Acute hemorrhagic cerebrovascular disease | White Race | - | Yes | 1.49 | 1.09 | 2.03 | 37550 | 8536 | 28673 | 45 | 296 | 0.005 | 0.010 | 0.005 | 201 |
| CIR021 | Acute hemorrhagic cerebrovascular disease | Black Race | - | - | 1.14 | 0.50 | 2.57 | 2881 | 648 | 2192 | * | 37 | * | 0.017 | * | * |
| CIR021 | Acute hemorrhagic cerebrovascular disease | Asian Race | - | - | 7.06 | 0.95 | 52.80 | 949 | 142 | 787 | * | 20 | * | 0.025 | * | * |
| CIR021 | Acute hemorrhagic cerebrovascular disease | Hispanic Race | - | - | 5.40 | 1.28 | 22.73 | 1968 | 397 | 1548 | * | 21 | * | 0.013 | * | * |
| CIR035 | Varicose veins of lower extremity | Overall | - | - | 1.19 | 0.86 | 1.65 | 44516 | 9934 | 34244 | 51 | 287 | 0.005 | 0.008 | 0.003 | 312 |
| CIR035 | Varicose veins of lower extremity | 1-60 days | - | Yes | 1.78 | 1.18 | 2.67 | 44516 | 9958 | 34373 | 27 | 158 | 0.003 | 0.005 | 0.002 | 534 |
| CIR035 | Varicose veins of lower extremity | 61-120 days | - | - | 0.81 | 0.46 | 1.43 | 27013 | 3631 | 23299 | 14 | 69 | 0.004 | 0.003 | -0.001 | n/a |
| CIR035 | Varicose veins of lower extremity | 121-182 days | - | - | 0.60 | 0.26 | 1.37 | 21622 | 2462 | 19090 | * | 60 | * | 0.003 | * | * |
| CIR035 | Varicose veins of lower extremity | Age 65-74 years | - | - | 0.99 | 0.60 | 1.65 | 20818 | 3995 | 16665 | 22 | 136 | 0.005 | 0.008 | 0.003 | 382 |
| CIR035 | Varicose veins of lower extremity | Age 75-84 years | - | - | 1.32 | 0.78 | 2.23 | 15131 | 3500 | 11515 | 18 | 98 | 0.005 | 0.008 | 0.003 | 301 |
| CIR035 | Varicose veins of lower extremity | Age 85+ years | - | - | 1.52 | 0.79 | 2.92 | 8567 | 2439 | 6064 | 11 | 53 | 0.004 | 0.009 | 0.004 | 240 |
| CIR035 | Varicose veins of lower extremity | Male | - | - | 1.32 | 0.84 | 2.09 | 21851 | 4545 | 17152 | 24 | 130 | 0.005 | 0.008 | 0.002 | 441 |
| CIR035 | Varicose veins of lower extremity | Female | - | - | 1.12 | 0.72 | 1.74 | 22665 | 5389 | 17092 | 27 | 157 | 0.005 | 0.009 | 0.004 | 243 |
| CIR035 | Varicose veins of lower extremity | White Race | - | - | 1.23 | 0.85 | 1.78 | 37597 | 8542 | 28768 | 43 | 244 | 0.005 | 0.008 | 0.003 | 294 |
| CIR035 | Varicose veins of lower extremity | Black Race | - | - | 0.58 | 0.18 | 1.91 | 2910 | 655 | 2245 | * | ≤10 | * | 0.003 | * | * |
| CIR035 | Varicose veins of lower extremity | Asian Race | - | - | 0.62 | 0.11 | 3.41 | 970 | 144 | 822 | * | ≤10 | * | 0.004 | * | * |
| CIR035 | Varicose veins of lower extremity | Hispanic Race | - | - | 1.69 | 0.48 | 5.98 | 1979 | 401 | 1548 | * | 28 | * | 0.018 | * | * |
| Abbreviations: A1 = Atorvastatin initiation; A2 = Newly initiated a different medication other than atorvastatin; N = Total number of patients in the analytic stratum; A = Number of A2 patients without the event; B = Number of A1 patients without the event; C = Number |  |  |  |  |  |  |  |  |  |  |  |  |  |  |  |  |

**Table S3 Continued: Weighted Fine-Gray and Descriptive Analyses for Trial 0 for Detected ADE Signals (Primary Analysis and Sensitivity Analysis)**
| Detected ADE Signals: Primary Algorithm ($q \leq 0.1$ & $sHR \geq 1.2$) and Sensitivity Analysis ($q \leq 0.2$ & $sHR \geq 1.2$) | | | | | | | | | | | | | | | | |
| --- | --- | --- | --- | --- | --- | --- | --- | --- | --- | --- | --- | --- | --- | --- | --- | --- |
| CCSR | CCSR Description | Strata | Detected ADE Signal Weighted Fine-Gray |  |  |  |  | Unadjusted Descriptive Analyses for Trial 0 |  |  |  |  |  |  |  |  |
|  |  |  | Primary | SA | sHR | LCL | UCL | N | A | B | C | D | Risk A2 | Risk A1 | RD | NNH |
| INJ024 | Sprains and strains, initial encounter | Overall | - | Yes | 1.34 | 1.10 | 1.63 | 43772 | 9665 | 33286 | 112 | 709 | 0.011 | 0.021 | 0.009 | 106 |
| INJ024 | Sprains and strains, initial encounter | 1-60 days | - | - | 1.22 | 0.94 | 1.59 | 43772 | 9713 | 33685 | 64 | 310 | 0.007 | 0.009 | 0.003 | 389 |
| INJ024 | Sprains and strains, initial encounter | 61-120 days | - | - | 1.76 | 1.17 | 2.66 | 26413 | 3513 | 22658 | 23 | 219 | 0.007 | 0.010 | 0.003 | 326 |
| INJ024 | Sprains and strains, initial encounter | 121-182 days | - | - | 1.06 | 0.68 | 1.65 | 21005 | 2363 | 18437 | 25 | 180 | 0.010 | 0.010 | -0.001 | n/a |
| INJ024 | Sprains and strains, initial encounter | Age 65-74 years | - | - | 1.38 | 1.02 | 1.87 | 20385 | 3872 | 16103 | 45 | 365 | 0.011 | 0.022 | 0.011 | 94 |
| INJ024 | Sprains and strains, initial encounter | Age 75-84 years | - | - | 1.52 | 1.07 | 2.15 | 14894 | 3403 | 11216 | 35 | 240 | 0.010 | 0.021 | 0.011 | 93 |
| INJ024 | Sprains and strains, initial encounter | Age 85+ years | - | - | 1.02 | 0.69 | 1.50 | 8493 | 2390 | 5967 | 32 | 104 | 0.013 | 0.017 | 0.004 | 255 |
| **INJ024** | **Sprains and strains, initial encounter** | **Male** | **Yes** | **Yes** | **1.79** | **1.26** | **2.54** | **21458** | **4443** | **16657** | **35** | **323** | **0.008** | **0.019** | **0.011** | **89** |
| INJ024 | Sprains and strains, initial encounter | Female | - | - | 1.14 | 0.90 | 1.45 | 22314 | 5222 | 16629 | 77 | 386 | 0.015 | 0.023 | 0.008 | 123 |
| INJ024 | Sprains and strains, initial encounter | White Race | - | - | 1.29 | 1.04 | 1.59 | 36946 | 8302 | 27937 | 103 | 604 | 0.012 | 0.021 | 0.009 | 112 |
| INJ024 | Sprains and strains, initial encounter | Black Race | - | - | 3.40 | 1.39 | 8.29 | 2850 | 643 | 2161 | * | 45 | * | 0.020 | * | * |
| INJ024 | Sprains and strains, initial encounter | Asian Race | - | - | 0.70 | 0.09 | 5.48 | 960 | 141 | 810 | * | * | * | * | * | * |
| INJ024 | Sprains and strains, initial encounter | Hispanic Race | - | - | 1.30 | 0.51 | 3.29 | 1962 | 388 | 1533 | * | 35 | * | 0.022 | * | * |
| SYM012 | Circulatory signs and symptoms | Overall | - | - | 1.06 | 0.99 | 1.13 | 34845 | 6426 | 21725 | 1214 | 5480 | 0.159 | 0.201 | 0.043 | 24 |
| SYM012 | Circulatory signs and symptoms | 1-60 days | - | - | 1.12 | 1.04 | 1.21 | 34845 | 6717 | 23298 | 923 | 3907 | 0.121 | 0.144 | 0.023 | 44 |
| SYM012 | Circulatory signs and symptoms | 61-120 days | - | - | 0.82 | 0.70 | 0.97 | 18015 | 2181 | 14664 | 187 | 983 | 0.079 | 0.063 | -0.016 | n/a |
| SYM012 | Circulatory signs and symptoms | 121-182 days | - | - | 0.82 | 0.66 | 1.02 | 13606 | 1400 | 11512 | 104 | 590 | 0.069 | 0.049 | -0.02 | n/a |
| SYM012 | Circulatory signs and symptoms | Age 65-74 years | - | - | 0.99 | 0.89 | 1.09 | 16515 | 2610 | 10718 | 501 | 2686 | 0.161 | 0.200 | 0.039 | 25 |
| SYM012 | Circulatory signs and symptoms | Age 75-84 years | - | - | 1.07 | 0.96 | 1.20 | 11706 | 2249 | 7167 | 422 | 1868 | 0.158 | 0.207 | 0.049 | 21 |
| SYM012 | Circulatory signs and symptoms | Age 85+ years | - | - | 1.19 | 1.03 | 1.37 | 6624 | 1567 | 3840 | 291 | 926 | 0.157 | 0.194 | 0.038 | 27 |
| SYM012 | Circulatory signs and symptoms | Male | - | - | 1.03 | 0.93 | 1.14 | 17128 | 3026 | 10969 | 537 | 2596 | 0.151 | 0.191 | 0.041 | 25 |
| SYM012 | Circulatory signs and symptoms | Female | - | - | 1.08 | 0.99 | 1.18 | 17717 | 3400 | 10756 | 677 | 2884 | 0.166 | 0.211 | 0.045 | 22 |
| CIR035 | Circulatory signs and symptoms | White Race | - | - | 1.02 | 0.95 | 1.10 | 29385 | 5506 | 18232 | 1058 | 4589 | 0.161 | 0.201 | 0.04 | 25 |
| **SYM012** | **Circulatory signs and symptoms** | **Black Race** | - | **Yes** | **1.51** | **1.16** | **1.96** | **2281** | **432** | **1415** | **66** | **368** | **0.133** | **0.206** | **0.074** | **14** |
| SYM012 | Circulatory signs and symptoms | Asian Race | - | - | 1.08 | 0.63 | 1.86 | 747 | 94 | 519 | 16 | 118 | 0.145 | 0.185 | 0.04 | 25 |
| SYM012 | Circulatory signs and symptoms | Hispanic Race | - | - | 1.37 | 1.01 | 1.87 | 1585 | 261 | 988 | 47 | 289 | 0.153 | 0.226 | 0.074 | 14 |

**Table S3 Continued: Weighted Fine-Gray and Descriptive Analyses for Trial 0 for Detected ADE Signals (Primary Analysis and Sensitivity Analysis)**
| Detected ADE Signals: Primary Algorithm ($q \leq 0.1$ & $sHR \geq 1.2$) and Sensitivity Analysis ($q \leq 0.2$ & $sHR \geq 1.2$) | | | | | | | | | | | | | | | | |
| --- | --- | --- | --- | --- | --- | --- | --- | --- | --- | --- | --- | --- | --- | --- | --- | --- |
| CCSR | CCSR Description | Strata | Detected ADE Signal Weighted Fine-Gray |  |  |  |  | Unadjusted Descriptive Analyses for Trial 0 |  |  |  |  |  |  |  |  |
|  |  |  | Primary | SA | sHR | LCL | UCL | N | A | B | C | D | Risk A2 | Risk A1 | RD | NNH |
| SYM014 | Skin/Subcutaneous signs and symptoms | Overall | - | - | 1.04 | 0.94 | 1.15 | 41168 | 8755 | 29313 | 518 | 2582 | 0.056 | 0.081 | 0.025 | 40 |
| SYM014 | Skin/Subcutaneous signs and symptoms | 1-60 days | - | - | 1.07 | 0.95 | 1.21 | 41168 | 8915 | 30352 | 358 | 1543 | 0.039 | 0.048 | 0.01 | 102 |
| SYM014 | Skin/Subcutaneous signs and symptoms | 61-120 days | - | - | 0.93 | 0.75 | 1.16 | 23821 | 3109 | 19999 | 104 | 609 | 0.032 | 0.030 | -0.003 | n/a |
| SYM014 | Skin/Subcutaneous signs and symptoms | 121-182 days | - | - | 0.86 | 0.64 | 1.15 | 18592 | 2046 | 16060 | 56 | 430 | 0.027 | 0.026 | -0.001 | n/a |
| SYM014 | Skin/Subcutaneous signs and symptoms | Age 65-74 years | - | - | 1.07 | 0.91 | 1.26 | 19179 | 3529 | 14216 | 188 | 1246 | 0.051 | 0.081 | 0.03 | 33 |
| SYM014 | Skin/Subcutaneous signs and symptoms | Age 75-84 years | - | - | 1.03 | 0.88 | 1.21 | 13948 | 3043 | 9796 | 198 | 911 | 0.061 | 0.085 | 0.024 | 42 |
| SYM014 | Skin/Subcutaneous signs and symptoms | Age 85+ years | - | - | 0.98 | 0.80 | 1.20 | 8041 | 2183 | 5301 | 132 | 425 | 0.057 | 0.074 | 0.017 | 58 |
| SYM014 | Skin/Subcutaneous signs and symptoms | Male | - | - | 1.04 | 0.89 | 1.22 | 20340 | 4068 | 14880 | 203 | 1189 | 0.048 | 0.074 | 0.026 | 38 |
| SYM014 | Skin/Subcutaneous signs and symptoms | Female | - | - | 1.04 | 0.92 | 1.19 | 20828 | 4687 | 14433 | 315 | 1393 | 0.063 | 0.088 | 0.025 | 40 |
| SYM014 | Skin/Subcutaneous signs and symptoms | White Race | - | - | 0.98 | 0.88 | 1.09 | 34770 | 7503 | 24635 | 465 | 2167 | 0.058 | 0.081 | 0.022 | 44 |
| SYM014 | Skin/Subcutaneous signs and symptoms | Black Race | - | Yes | 1.86 | 1.22 | 2.84 | 2664 | 581 | 1888 | 24 | 171 | 0.040 | 0.083 | 0.043 | 23 |
| SYM014 | Skin/Subcutaneous signs and symptoms | Asian Race | - | - | 1.78 | 0.77 | 4.12 | 887 | 130 | 680 | * | * | * | * | * | * |
| SYM014 | Skin/Subcutaneous signs and symptoms | Hispanic Race | - | - | 0.91 | 0.56 | 1.47 | 1841 | 364 | 1357 | 17 | 103 | 0.045 | 0.071 | 0.026 | 39 |
| SYM015 | General sensation/perception signs and symptoms | Overall | Yes | Yes | 1.23 | 1.11 | 1.36 | 40054 | 8394 | 28368 | 526 | 2766 | 0.059 | 0.089 | 0.030 | 33 |
| SYM015 | General sensation/perception signs and symptoms | 1-60 days | - | Yes | 1.20 | 1.07 | 1.36 | 40054 | 8531 | 29393 | 389 | 1741 | 0.044 | 0.056 | 0.012 | 81 |
| SYM015 | General sensation/perception signs and symptoms | 61-120 days | - | - | 1.24 | 0.98 | 1.56 | 22996 | 2965 | 19301 | 85 | 645 | 0.028 | 0.032 | 0.004 | 224 |
| SYM015 | General sensation/perception signs and symptoms | 121-182 days | - | - | 1.16 | 0.87 | 1.56 | 17911 | 1961 | 15518 | 52 | 380 | 0.026 | 0.024 | -0.002 | n/a |
| SYM015 | General sensation/perception signs and symptoms | Age 65-74 years | - | - | 1.16 | 0.99 | 1.36 | 19186 | 3496 | 14183 | 205 | 1302 | 0.055 | 0.084 | 0.029 | 35 |
| SYM015 | General sensation/perception signs and symptoms | Age 75-84 years | - | - | 1.21 | 1.03 | 1.42 | 13404 | 2879 | 9344 | 203 | 978 | 0.066 | 0.095 | 0.029 | 35 |
| SYM015 | General sensation/perception signs and symptoms | Age 85+ years | - | Yes | 1.46 | 1.18 | 1.81 | 7464 | 2019 | 4841 | 118 | 486 | 0.055 | 0.091 | 0.036 | 28 |
| SYM015 | General sensation/perception signs and symptoms | Male | - | Yes | 1.24 | 1.06 | 1.44 | 19985 | 3918 | 14518 | 237 | 1312 | 0.057 | 0.083 | 0.026 | 39 |
| SYM015 | General sensation/perception signs and symptoms | Female | - | - | 1.23 | 1.07 | 1.40 | 20069 | 4476 | 13850 | 289 | 1454 | 0.061 | 0.095 | 0.034 | 29 |
| SYM015 | General sensation/perception signs and symptoms | White Race | Yes | Yes | 1.20 | 1.08 | 1.34 | 33902 | 7244 | 23888 | 456 | 2314 | 0.059 | 0.088 | 0.029 | 34 |
| SYM015 | General sensation/perception signs and symptoms | Black Race | - | Yes | 1.76 | 1.18 | 2.63 | 2568 | 549 | 1822 | 32 | 165 | 0.055 | 0.083 | 0.028 | 36 |
| SYM015 | General sensation/perception signs and symptoms | Asian Race | - | - | 1.48 | 0.70 | 3.11 | 854 | 116 | 663 | * | 70 | * | 0.095 | * | * |
| SYM015 | General sensation/perception signs and symptoms | Hispanic Race | - | - | 0.90 | 0.54 | 1.49 | 1757 | 317 | 1279 | 26 | 135 | 0.076 | 0.095 | 0.020 | 51 |

**Table S3 Continued: Weighted Fine-Gray and Descriptive Analyses for Trial 0 for Detected ADE Signals (Primary Analysis and Sensitivity Analysis)**
| Detected ADE Signals: Primary Algorithm (q ≤ 0.1 & sHR ≥ 1.2) and Sensitivity Analysis (q ≤ 0.2 & sHR ≥ 1.2) |  |  |  |  |  |  |  |  |  |  |  |  |  |  |  |  |
| --- | --- | --- | --- | --- | --- | --- | --- | --- | --- | --- | --- | --- | --- | --- | --- | --- |
| CCSR | CCSR Description | Strata | Detected ADE Signal Weighted Fine-Gray |  |  |  |  | Unadjusted Descriptive Analyses for Trial 0 |  |  |  |  |  |  |  |  |
|  |  |  | Primary | SA | sHR | LCL | UCL | N | A | B | C | D | Risk A2 | Risk A1 | RD | NNH |
| SYM017 | Abnormal findings without diagnosis | Overall | - | - | 1.07 | 0.99 | 1.14 | 24345 | 3637 | 13008 | 1278 | 6422 | 0.260 | 0.331 | 0.070 | 14 |
| SYM017 | Abnormal findings without diagnosis | 1-60 days | - | - | 1.02 | 0.94 | 1.10 | 24345 | 3865 | 14932 | 1050 | 4498 | 0.214 | 0.231 | 0.018 | 56 |
| SYM017 | Abnormal findings without diagnosis | 61-120 days | - | - | 1.12 | 0.93 | 1.34 | 11527 | 1212 | 8922 | 163 | 1230 | 0.119 | 0.121 | 0.003 | 383 |
| **SYM017** | **Abnormal findings without diagnosis** | **121-182 days** | **Yes** | - | **1.55** | **1.18** | **2.05** | **8269** | **820** | **6690** | **65** | **694** | **0.073** | **0.094** | **0.021** | **49** |
| SYM017 | Abnormal findings without diagnosis | Age 65-74 years | - | - | 0.99 | 0.89 | 1.10 | 11591 | 1478 | 6463 | 513 | 3137 | 0.258 | 0.327 | 0.069 | 14 |
| SYM017 | Abnormal findings without diagnosis | Age 75-84 years | - | - | 1.13 | 1.00 | 1.27 | 8047 | 1240 | 4195 | 439 | 2173 | 0.261 | 0.341 | 0.080 | 13 |
| SYM017 | Abnormal findings without diagnosis | Age 85+ years | - | - | 1.13 | 0.99 | 1.29 | 4707 | 919 | 2350 | 326 | 1112 | 0.262 | 0.321 | 0.059 | 17 |
| SYM017 | Abnormal findings without diagnosis | Male | - | - | 1.03 | 0.93 | 1.14 | 11740 | 1656 | 6368 | 572 | 3144 | 0.257 | 0.331 | 0.074 | 14 |
| SYM017 | Abnormal findings without diagnosis | Female | - | - | 1.10 | 1.00 | 1.20 | 12605 | 1981 | 6640 | 706 | 3278 | 0.263 | 0.331 | 0.068 | 15 |
| SYM017 | Abnormal findings without diagnosis | White Race | - | - | 1.08 | 1.00 | 1.16 | 20511 | 3132 | 10908 | 1089 | 5382 | 0.258 | 0.330 | 0.072 | 14 |
| SYM017 | Abnormal findings without diagnosis | Black Race | - | - | 0.95 | 0.71 | 1.27 | 1618 | 232 | 887 | 86 | 413 | 0.270 | 0.318 | 0.047 | 21 |
| SYM017 | Abnormal findings without diagnosis | Asian Race | - | - | 2.46 | 1.30 | 4.67 | 527 | 58 | 288 | 13 | 168 | 0.183 | 0.368 | 0.185 | 5 |
| SYM017 | Abnormal findings without diagnosis | Hispanic Race | - | - | 1.06 | 0.77 | 1.47 | 1107 | 147 | 596 | 55 | 309 | 0.272 | 0.341 | 0.069 | 14 |
| SYM018 | Prediabetes | Overall | - | - | 1.48 | 1.08 | 2.03 | 43707 | 9871 | 33278 | 52 | 506 | 0.005 | 0.015 | 0.010 | 103 |
| **SYM018** | **Prediabetes** | **1-60 days** | - | **Yes** | **1.64** | **1.11** | **2.40** | **43707** | **9889** | **33487** | **34** | **297** | **0.003** | **0.009** | **0.005** | **186** |
| SYM018 | Prediabetes | 61-120 days | - | - | 1.06 | 0.52 | 2.15 | 26312 | 3608 | 22568 | 13 | 123 | 0.004 | 0.005 | 0.002 | 546 |
| SYM018 | Prediabetes | 121-182 days | - | - | 1.24 | 0.54 | 2.84 | 21010 | 2446 | 18473 | * | 86 | * | 0.005 | * | * |
| SYM018 | Prediabetes | Age 65-74 years | - | - | 1.31 | 0.80 | 2.14 | 20268 | 3948 | 16016 | 20 | 284 | 0.005 | 0.017 | 0.012 | 81 |
| SYM018 | Prediabetes | Age 75-84 years | - | - | 1.80 | 1.15 | 2.82 | 14923 | 3477 | 11259 | 24 | 163 | 0.007 | 0.014 | 0.007 | 135 |
| SYM018 | Prediabetes | Age 85+ years | - | - | 1.33 | 0.72 | 2.46 | 8516 | 2446 | 6003 | * | 59 | * | 0.010 | * | * |
| SYM018 | Prediabetes | Male | - | - | 1.50 | 0.94 | 2.39 | 21312 | 4503 | 16512 | 25 | 272 | 0.006 | 0.016 | 0.011 | 94 |
| SYM018 | Prediabetes | Female | - | - | 1.45 | 0.94 | 2.22 | 22395 | 5368 | 16766 | 27 | 234 | 0.005 | 0.014 | 0.009 | 114 |
| **SYM018** | **Prediabetes** | **White Race** | **Yes** | **Yes** | **1.71** | **1.24** | **2.36** | **36982** | **8498** | **28052** | **43** | **389** | **0.005** | **0.014** | **0.009** | **116** |
| SYM018 | Prediabetes | Black Race | - | - | 1.81 | 0.49 | 6.61 | 2826 | 647 | 2120 | * | 55 | * | 0.025 | * | * |
| SYM018 | Prediabetes | Asian Race | - | - | 0.27 | 0.07 | 1.10 | 932 | 135 | 781 | * | 14 | * | 0.018 | * | * |
| SYM018 | Prediabetes | Hispanic Race | - | - | 0.77 | 0.26 | 2.25 | 1932 | 401 | 1500 | * | 29 | * | 0.019 | * | * |
| Abbreviations: A1 = Atorvastatin initiation; A2 = Newly initiated a different medication other than atorvastatin; N = Total number of patients in the analytic stratum; A = Number of A2 patients without the event; B = Number of A1 patients without the event; C = Number of A2 patients with the event; D = Number of A1 patients with the event; Risk A1 = Cumulative incidence (risk) in the A1 (atorvastatin) group; Risk A2 = Cumulative incidence (risk) in the A2 (comparator) group; RD = Risk difference; NNH = Number needed to harm; Primary = ADE signal detected using the primary ADE signal detection algorithm (q<=0.1 & sHR >=1.2); SA = ADE signal detected in the sensitivity analysis using the ADE signal detection algorithm (q<=0.2 & sHR >=1.2); * = Not reportable due to cell counts ≤ 10 per CMS policies |  |  |  |  |  |  |  |  |  |  |  |  |  |  |  |  |

**Table S4.** Frequency Distribution of Observed ICD-10-CM Codes Comprising Each Detected ADE Signal During Follow-Up.

| CCSR | ICD-10-CM | ICD-10-CM Code Description | Freq | Percent |
| --- | --- | --- | --- | --- |
| BLD004 | D62 | Acute posthemorrhagic anemia | 3238 | 100 |
| CIR003 | I340 | Nonrheumatic mitral (valve) insufficiency | 5996 | 45.99 |
| CIR003 | I350 | Nonrheumatic aortic (valve) stenosis | 2700 | 20.71 |
| CIR003 | I351 | Nonrheumatic aortic (valve) insufficiency | 1197 | 9.18 |
| CIR003 | Z952 | Presence of prosthetic heart valve | 659 | 5.05 |
| CIR003 | I361 | Nonrheumatic tricuspid (valve) insufficiency | 609 | 4.67 |
| CIR003 | I358 | Other nonrheumatic aortic valve disorders | 400 | 3.07 |
| CIR003 | I341 | Nonrheumatic mitral (valve) prolapse | 327 | 2.51 |
| CIR003 | I348 | Other nonrheumatic mitral valve disorders | 308 | 2.36 |
| CIR003 | I359 | Nonrheumatic aortic valve disorder, unspecified | 229 | 1.76 |
| CIR003 | I371 | Nonrheumatic pulmonary valve insufficiency | 129 | 0.99 |
| CIR003 | I342 | Nonrheumatic mitral (valve) stenosis | 96 | 0.74 |
| CIR003 | I352 | Nonrheumatic aortic (valve) stenosis with insuffic | 83 | 0.64 |
| CIR003 | I349 | Nonrheumatic mitral valve disorder, unspecified | 72 | 0.55 |
| CIR003 | Z953 | Presence of xenogenic heart valve | 66 | 0.51 |
| CIR003 | Z954 | Presence of other heart-valve replacement | 63 | 0.48 |
| CIR003 | I368 | Other nonrheumatic tricuspid valve disorders | 35 | 0.27 |
| CIR003 | I360 | Nonrheumatic tricuspid (valve) stenosis | 29 | 0.22 |
| CIR003 | I369 | Nonrheumatic tricuspid valve disorder, unspecified | 27 | 0.21 |
| CIR021 | I619 | Nontraumatic intracerebral hemorrhage, unspecified | 277 | 24.41 |
| CIR021 | I609 | Nontraumatic subarachnoid hemorrhage, unspecified | 168 | 14.8 |
| CIR021 | I6200 | Nontraumatic subdural hemorrhage, unspecified | 160 | 14.1 |
| CIR021 | I629 | Nontraumatic intracranial hemorrhage, unspecified | 157 | 13.83 |
| CIR021 | I611 | Nontraumatic intracerebral hemorrhage in hemispher | 75 | 6.61 |
| CIR021 | I618 | Other nontraumatic intracerebral hemorrhage | 73 | 6.43 |
| CIR021 | I610 | Nontraumatic intracerebral hemorrhage in hemispher | 40 | 3.52 |
| CIR021 | I614 | Nontraumatic intracerebral hemorrhage in cerebellu | 29 | 2.56 |
| CIR021 | I615 | Nontraumatic intracerebral hemorrhage, intraventri | 27 | 2.38 |
| CIR021 | I6203 | Nontraumatic chronic subdural hemorrhage | 27 | 2.38 |
| CIR021 | I613 | Nontraumatic intracerebral hemorrhage in brain ste | 25 | 2.2 |
| CIR035 | I8393 | Asymptomatic varicose veins of bilateral lower ext | 133 | 13.35 |
| CIR035 | I83893 | Varicose veins of bilateral lower extremities with | 131 | 13.15 |
| CIR035 | I8390 | Asymptomatic varicose veins of unspecified lower e | 119 | 11.95 |
| CIR035 | I83813 | Varicose veins of bilateral lower extremities with | 99 | 9.94 |
| CIR035 | I8310 | Varicose veins of unspecified lower extremity with | 87 | 8.73 |
| CIR035 | I8311 | Varicose veins of right lower extremity with infla | 83 | 8.33 |
| CIR035 | I83009 | Varicose veins of unspecified lower extremity with | 35 | 3.51 |
| CIR035 | I83899 | Varicose veins of unspecified lower extremity with | 30 | 3.01 |
| CIR035 | I8312 | Varicose veins of left lower extremity with inflam | 24 | 2.41 |
| CIR035 | I83018 | Varicose veins of right lower extremity with ulcer | 22 | 2.21 |
| CIR035 | I83891 | Varicose veins of right lower extremity with other | 22 | 2.21 |
| INJ024 | S161XXA | Strain of muscle, fascia and tendon at neck level, | 208 | 10.01 |
| INJ024 | S39012A | Strain of muscle, fascia and tendon of lower back, | 149 | 7.17 |
| INJ024 | S134XXA | Sprain of ligaments of cervical spine, initial enc | 116 | 5.58 |
| INJ024 | S335XXA | Sprain of ligaments of lumbar spine, initial encou | 111 | 5.34 |
| INJ024 | S93401A | Sprain of unspecified ligament of right ankle, ini | 75 | 3.61 |
| INJ024 | S336XXA | Sprain of sacroiliac joint, initial encounter | 67 | 3.23 |
| INJ024 | S233XXA | Sprain of ligaments of thoracic spine, initial enc | 66 | 3.18 |
| INJ024 | S93402A | Sprain of unspecified ligament of left ankle, init | 64 | 3.08 |
| INJ024 | S46011A | Strain of muscle(s) and tendon(s) of the rotator c | 47 | 2.26 |
| INJ024 | S63502A | Unspecified sprain of left wrist, initial encounte | 38 | 1.83 |
| INJ024 | S29012A | Strain of muscle and tendon of back wall of thorax | 38 | 1.83 |
| INJ024 | S338XXA | Sprain of other parts of lumbar spine and pelvis, | 36 | 1.73 |
| INJ024 | S46912A | Strain of unspecified muscle, fascia and tendon at | 30 | 1.44 |
| INJ024 | S29011A | Strain of muscle and tendon of front wall of thora | 29 | 1.4 |
| INJ024 | S63501A | Unspecified sprain of right wrist, initial encount | 29 | 1.4 |
| INJ024 | S83241A | Other tear of medial meniscus, current injury, rig | 29 | 1.4 |
| INJ024 | S46911A | Strain of unspecified muscle, fascia and tendon at | 29 | 1.4 |
| INJ024 | S46811A | Strain of other muscles, fascia and tendons at sho | 28 | 1.35 |
| INJ024 | S76011A | Strain of muscle, fascia and tendon of right hip, | 25 | 1.2 |
| INJ024 | S83242A | Other tear of medial meniscus, current injury, lef | 25 | 1.2 |
| INJ024 | S43401A | Unspecified sprain of right shoulder joint, initia | 24 | 1.16 |
| INJ024 | S46012A | Strain of muscle(s) and tendon(s) of the rotator c | 23 | 1.11 |
| INJ024 | S93601A | Unspecified sprain of right foot, initial encounte | 22 | 1.06 |
| INJ024 | S8392XA | Sprain of unspecified site of left knee, initial e | 21 | 1.01 |
| INJ024 | S39011A | Strain of muscle, fascia and tendon of abdomen, in | 21 | 1.01 |
| INJ024 | S2341XA | Sprain of ribs, initial encounter | 20 | 0.96 |
| INJ024 | S46812A | Strain of other muscles, fascia and tendons at sho | 20 | 0.96 |
| INJ024 | S83231A | Complex tear of medial meniscus, current injury, r | 20 | 0.96 |
| SYM012 | R001 | Bradycardia, unspecified | 5210 | 29.52 |
| SYM012 | R002 | Palpitations | 3237 | 18.34 |
| SYM012 | R0989 | Other specified symptoms and signs involving the c | 3199 | 18.13 |
| SYM012 | R000 | Tachycardia, unspecified | 2883 | 16.34 |
| SYM012 | R030 | Elevated blood-pressure reading, without diagnosis | 1239 | 7.02 |
| SYM012 | R011 | Cardiac murmur, unspecified | 902 | 5.11 |
| SYM012 | R58 | Hemorrhage, not elsewhere classified | 582 | 3.3 |
| SYM012 | R031 | Nonspecific low blood-pressure reading | 209 | 1.18 |
| SYM012 | R008 | Other abnormalities of heart beat | 113 | 0.64 |
| SYM012 | R010 | Benign and innocent cardiac murmurs | 34 | 0.19 |
| SYM012 | R009 | Unspecified abnormalities of heart beat | 24 | 0.14 |
| SYM014 | R21 | Rash and other nonspecific skin eruption | 1420 | 18.56 |
| SYM014 | R200 | Anesthesia of skin | 1236 | 16.15 |
| SYM014 | R202 | Paresthesia of skin | 1103 | 14.41 |
| SYM014 | R220 | Localized swelling, mass and lump, head | 381 | 4.98 |
| SYM014 | R208 | Other disturbances of skin sensation | 368 | 4.81 |
| SYM014 | R238 | Other skin changes | 360 | 4.7 |
| SYM014 | R233 | Spontaneous ecchymoses | 316 | 4.13 |
| SYM014 | R2241 | Localized swelling, mass and lump, right lower lim | 275 | 3.59 |
| SYM014 | R221 | Localized swelling, mass and lump, neck | 269 | 3.52 |
| SYM014 | R222 | Localized swelling, mass and lump, trunk | 258 | 3.37 |
| SYM014 | R2243 | Localized swelling, mass and lump, lower limb, bil | 248 | 3.24 |
| SYM014 | R2242 | Localized swelling, mass and lump, left lower limb | 231 | 3.02 |
| SYM014 | R209 | Unspecified disturbances of skin sensation | 194 | 2.54 |
| SYM014 | R2231 | Localized swelling, mass and lump, right upper lim | 158 | 2.06 |
| SYM014 | R2232 | Localized swelling, mass and lump, left upper limb | 147 | 1.92 |
| SYM014 | R234 | Changes in skin texture | 135 | 1.76 |
| SYM014 | R231 | Pallor | 133 | 1.74 |
| SYM014 | R229 | Localized swelling, mass and lump, unspecified | 107 | 1.4 |
| SYM014 | R232 | Flushing | 78 | 1.02 |
| SYM014 | R239 | Unspecified skin changes | 63 | 0.82 |
| SYM014 | R230 | Cyanosis | 50 | 0.65 |
| SYM014 | R2240 | Localized swelling, mass and lump, unspecified low | 44 | 0.58 |
| SYM014 | R201 | Hypoesthesia of skin | 32 | 0.42 |
| SYM014 | R203 | Hyperesthesia | 25 | 0.33 |
| SYM015 | R42 | Dizziness and giddiness | 8120 | 95.78 |
| SYM015 | R443 | Hallucinations, unspecified | 175 | 2.06 |
| SYM015 | R441 | Visual hallucinations | 117 | 1.38 |
| SYM015 | R440 | Auditory hallucinations | 32 | 0.38 |
| SYM017 | R9431 | Abnormal electrocardiogram [ECG] [EKG] | 6621 | 22.13 |
| SYM017 | R918 | Other nonspecific abnormal finding of lung field | 4409 | 14.73 |
| SYM017 | R7989 | Other specified abnormal findings of blood chemist | 2569 | 8.58 |
| SYM017 | R748 | Abnormal levels of other serum enzymes | 1899 | 6.35 |
| SYM017 | R739 | Hyperglycemia, unspecified | 1777 | 5.94 |
| SYM017 | R911 | Solitary pulmonary nodule | 1208 | 4.04 |
| SYM017 | R7309 | Other abnormal glucose | 1189 | 3.97 |
| SYM017 | R7301 | Impaired fasting glucose | 1155 | 3.86 |
| SYM017 | R9720 | Elevated prostate specific antigen [PSA] | 965 | 3.22 |
| SYM017 | R791 | Abnormal coagulation profile | 650 | 2.17 |
| SYM017 | R9439 | Abnormal result of other cardiovascular function s | 485 | 1.62 |
| SYM017 | R945 | Abnormal results of liver function studies | 446 | 1.49 |
| SYM017 | R740 | Nonspecific elevation of levels of transaminase an | 444 | 1.48 |
| SYM017 | R799 | Abnormal finding of blood chemistry, unspecified | 414 | 1.38 |
| SYM017 | R931 | Abnormal findings on diagnostic imaging of heart a | 376 | 1.26 |
| SYM017 | R9082 | White matter disease, unspecified | 323 | 1.08 |
| SYM017 | R7881 | Bacteremia | 273 | 0.91 |
| SYM017 | R7302 | Impaired glucose tolerance (oral) | 270 | 0.9 |
| SYM017 | R9430 | Abnormal result of cardiovascular function study, | 251 | 0.84 |
| SYM017 | R946 | Abnormal results of thyroid function studies | 250 | 0.84 |
| SYM017 | R9089 | Other abnormal findings on diagnostic imaging of c | 247 | 0.83 |
| SYM017 | R930 | Abnormal findings on diagnostic imaging of skull a | 246 | 0.82 |
| SYM017 | R938 | Abnormal findings on diagnostic imaging of other s | 234 | 0.78 |
| SYM017 | R778 | Other specified abnormalities of plasma proteins | 230 | 0.77 |
| SYM017 | R928 | Other abnormal and inconclusive findings on diagno | 227 | 0.76 |
| SYM017 | R9389 | Abnormal findings on diagnostic imaging of other s | 183 | 0.61 |
| SYM017 | R944 | Abnormal results of kidney function studies | 179 | 0.6 |
| SYM017 | R921 | Mammographic calcification found on diagnostic ima | 160 | 0.53 |
| SYM017 | R935 | Abnormal findings on diagnostic imaging of other a | 152 | 0.51 |
| SYM017 | R933 | Abnormal findings on diagnostic imaging of other p | 151 | 0.5 |
| SYM017 | R768 | Other specified abnormal immunological findings in | 129 | 0.43 |
| SYM017 | R937 | Abnormal findings on diagnostic imaging of other p | 123 | 0.41 |
| SYM017 | R922 | Inconclusive mammogram | 108 | 0.36 |
| SYM017 | R899 | Unspecified abnormal finding in specimens from oth | 105 | 0.35 |
| SYM017 | R700 | Elevated erythrocyte sedimentation rate | 104 | 0.35 |
| SYM017 | R7889 | Finding of other specified substances, not normall | 90 | 0.3 |
| SYM017 | R942 | Abnormal results of pulmonary function studies | 85 | 0.28 |
| SYM017 | R790 | Abnormal level of blood mineral | 84 | 0.28 |
| SYM017 | R718 | Other abnormality of red blood cells | 76 | 0.25 |
| SYM017 | R7982 | Elevated C-reactive protein (CRP) | 75 | 0.25 |
| SYM017 | R7981 | Abnormal blood-gas level | 67 | 0.22 |
| SYM017 | R932 | Abnormal findings on diagnostic imaging of liver a | 65 | 0.22 |
| SYM017 | R9401 | Abnormal electroencephalogram [EEG] | 56 | 0.19 |
| SYM017 | R710 | Precipitous drop in hematocrit | 55 | 0.18 |
| SYM017 | R9341 | Abnormal radiologic findings on diagnostic imaging | 48 | 0.16 |
| SYM017 | R760 | Raised antibody titer | 46 | 0.15 |
| SYM017 | R9721 | Rising PSA following treatment for malignant neopl | 44 | 0.15 |
| SYM017 | R978 | Other abnormal tumor markers | 43 | 0.14 |
| SYM017 | R749 | Abnormal serum enzyme level, unspecified | 36 | 0.12 |
| SYM017 | R936 | Abnormal findings on diagnostic imaging of limbs | 35 | 0.12 |
| SYM017 | R7611 | Nonspecific reaction to tuberculin skin test witho | 34 | 0.11 |
| SYM017 | R9402 | Abnormal brain scan | 32 | 0.11 |
| SYM017 | R920 | Mammographic microcalcification found on diagnosti | 23 | 0.08 |
| SYM017 | R970 | Elevated carcinoembryonic antigen [CEA] | 22 | 0.07 |
| SYM017 | R770 | Abnormality of albumin | 22 | 0.07 |
| SYM017 | R81 | Glycosuria | 21 | 0.07 |
| SYM018 | R7303 | Prediabetes | 1581 | 100 |

**Table S5.**
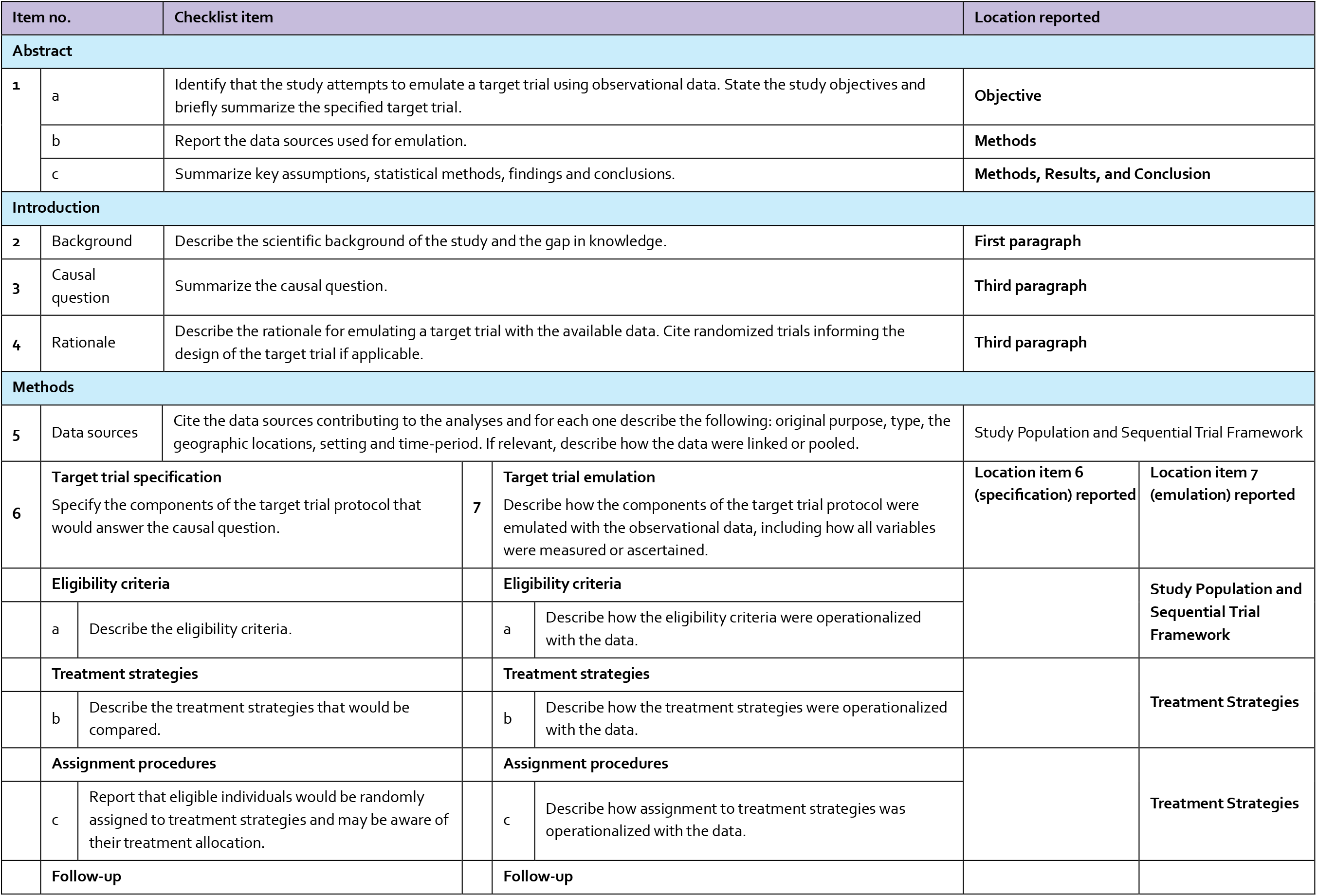
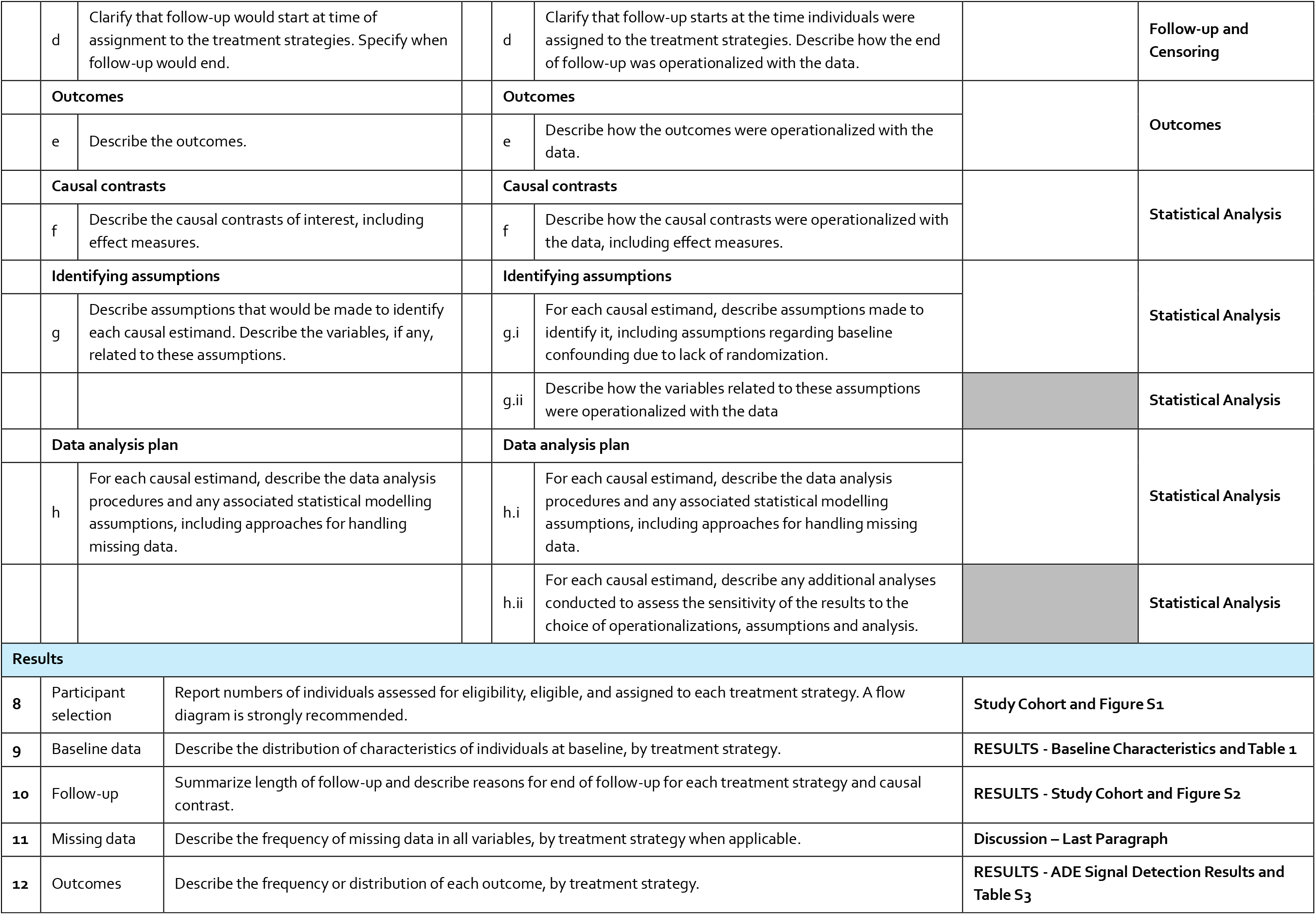
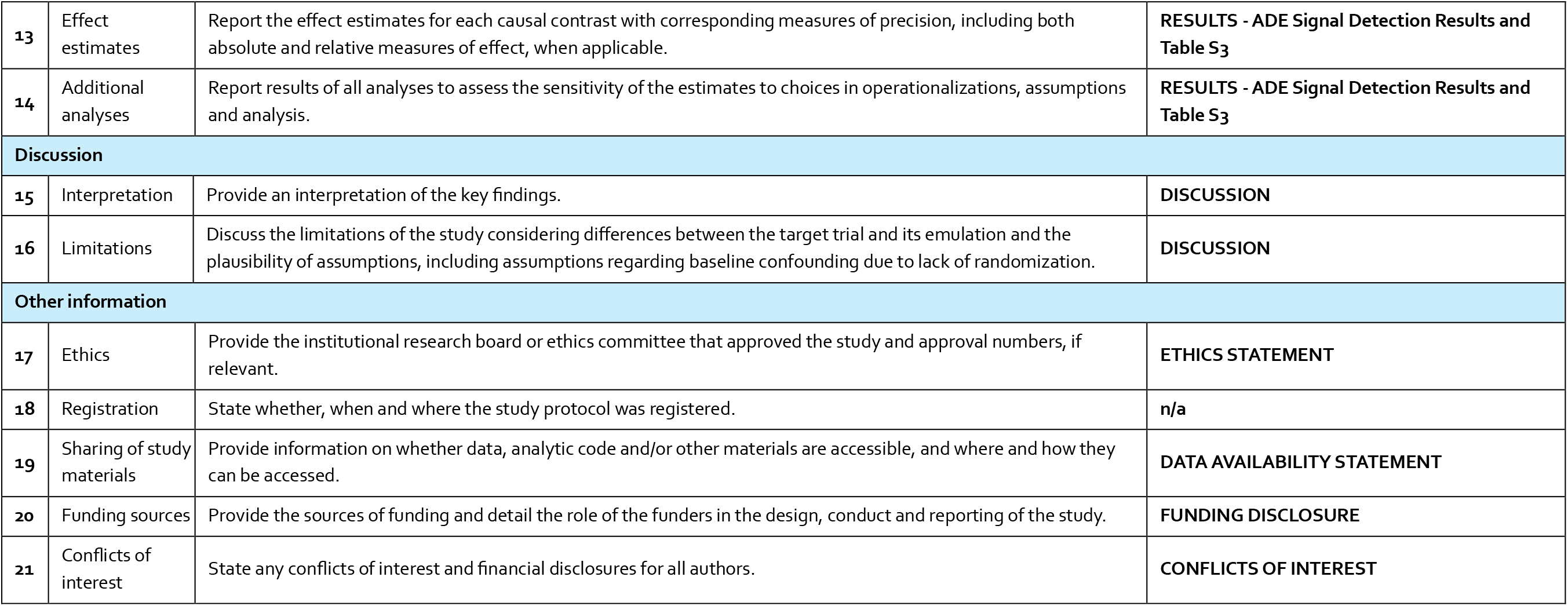
TARGET Checklist.

**Table S6.**
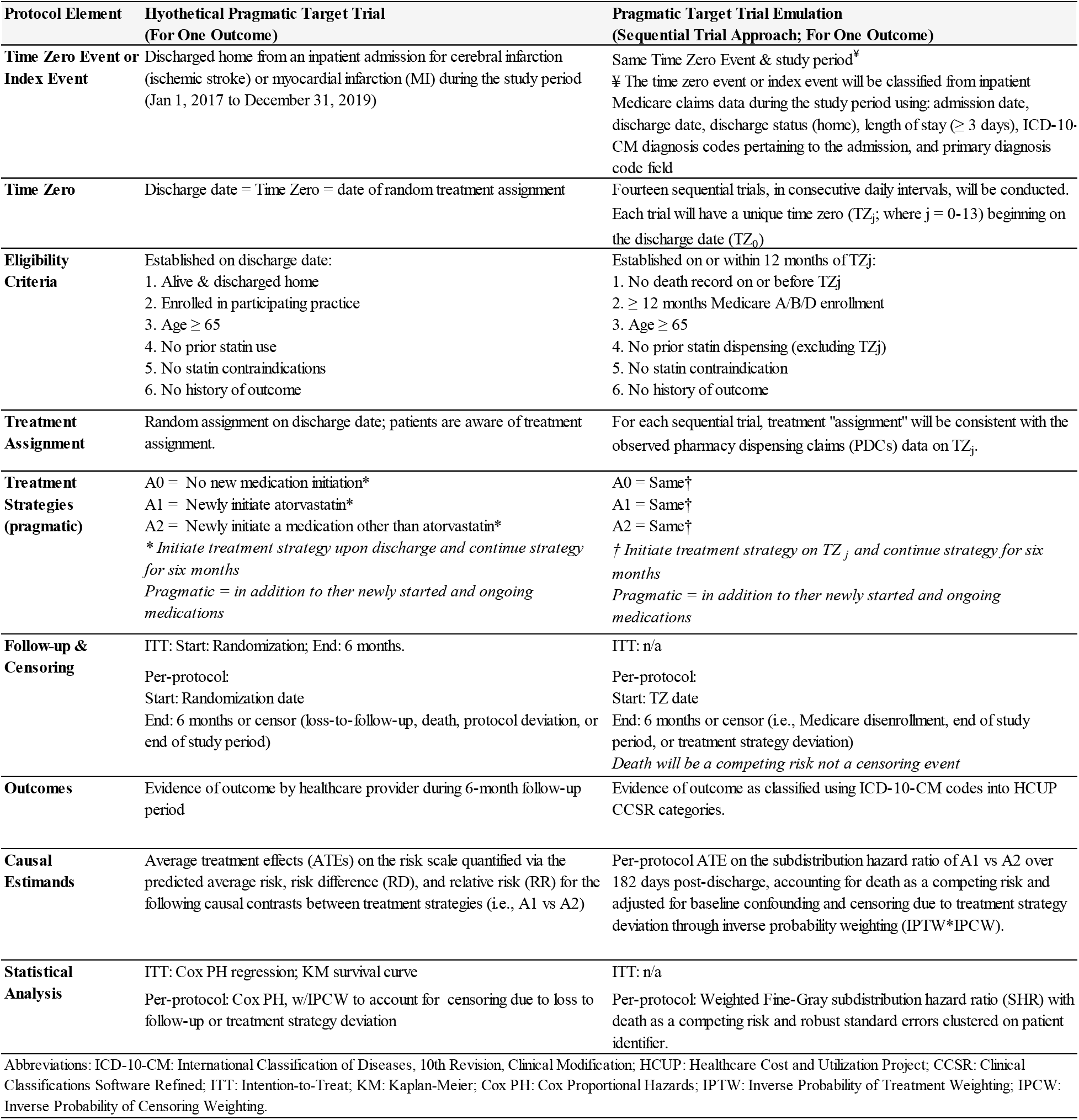
Protocol Elements of the Hypothetical Pragmatic Target Trial and Its Emulation Using a Sequential Trial Approach (For One Outcome)

**Figure S1.**
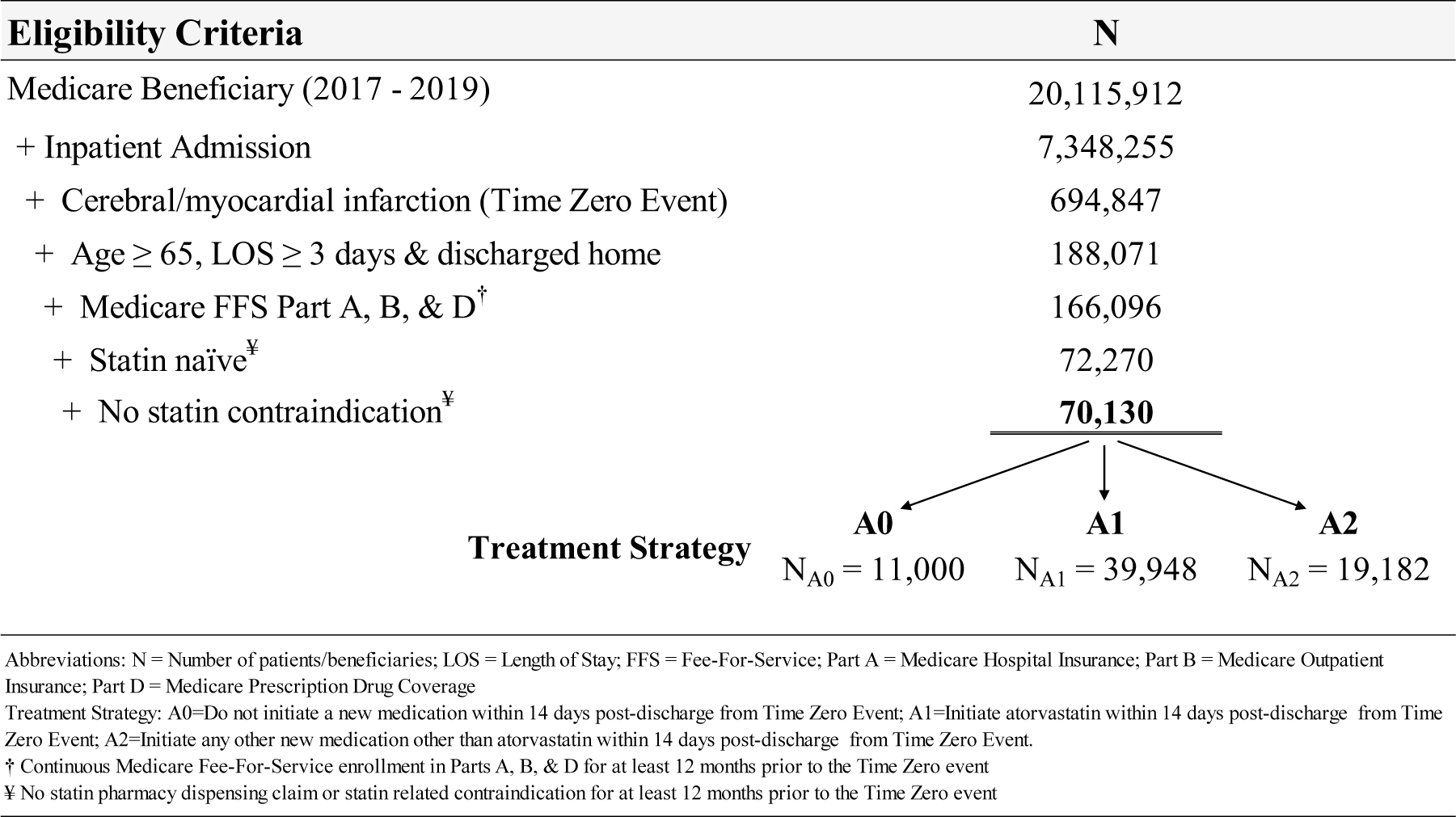
Eligibility Disposition and Treatment Strategy Initiation during the 14-Day Trial Period.

